# Effectiveness of nirmatrelvir-ritonavir against hospital admission or death: a cohort study in a large US healthcare system

**DOI:** 10.1101/2022.10.02.22280623

**Authors:** Joseph A. Lewnard, John M. McLaughlin, Debbie Malden, Vennis Hong, Laura Puzniak, Bradley K. Ackerson, Bruno J. Lewin, Jeniffer S. Kim, Sally F. Shaw, Harpreet Takhar, Luis Jodar, Sara Y. Tartof

## Abstract

**Background:** In the United States, oral nirmatrelvir-ritonavir (Paxlovid™) is authorized for use among patients aged ≥12 years with mild-to-moderate SARS-CoV-2 infection who are at risk for progression to severe COVID-19, including hospitalization. However, effectiveness under current real-world prescribing practices in outpatient settings is unclear.

**Methods:** We undertook a matched observational cohort study of non-hospitalized cases with SARS-CoV-2 infection to compare outcomes among those who received or did not receive nirmatrelvir-ritonavir within the Kaiser Permanente Southern California healthcare system. Cases were matched on testing date, age, sex, clinical status (including care received, presence or absence of acute COVID-19 symptoms at testing, and time from symptom onset to testing), history of vaccination, Charlson comorbidity index, prior-year healthcare utilization, and body mass index. Primary analyses evaluated effectiveness of nirmatrelvir-ritonavir in preventing hospital admission or death within 30 days after a positive test. Secondary analyses evaluated effectiveness against intensive care unit admission, mechanical ventilation, or death within 60 days after a positive test. We measured treatment effectiveness as (1–adjusted hazards ratio [aHR])×100%, estimating the aHR via Cox proportional hazards models.

**Results:** Analyses included 7,274 nirmatrelvir-ritonavir recipients and 126,152 non-recipients with positive results from SARS-CoV-2 tests undertaken in outpatient settings between 8 April and 7 October, 2022. Overall, 114,208 (85.6%) and 81,739 (61.3%) of 133,426 participants had received ≥2 and ≥3 COVID-19 vaccine doses, respectively. A total of 111,489 (83.6% of 133,426) cases were symptomatic at the point of testing, with 5,472 (75.2% of 7,274) treatment recipients and 84,657 (67.1% of 126,152) non-recipients testing within 0–5 days after symptom onset. Effectiveness in preventing hospital admission or death within 30 days after a positive test was 79.6% (95% confidence interval: 33.9% to 93.8%) for cases dispensed nirmatrelvir-ritonavir within 0–5 days after symptom onset; within the subgroup of cases tested 0–5 days after symptom onset and dispensed treatment on the day of their test, effectiveness was 89.6% (50.2% to 97.8%).

Effectiveness declined to 43.8% (–33.3% to 81.7%) for treatment course dispensed ≥6 days after symptom onset or to cases who were not experiencing acute clinical symptoms. Overall, for cases dispensed treatment at any time within their clinical course, effectiveness was 53.6% (6.6% to 77.0%). Effectiveness in preventing the secondary endpoint of intensive care unit admission, mechanical ventilation, or death within 60 days after a positive test was 89.2% (–25.0% to 99.3%) for cases dispensed treatment 0–5 days after symptom onset and 84.1% (18.8% to 96.9%) for cases dispensed treatment at any time. Subgroup analyses identified similar effectiveness estimates among cases who had received ≥2 or ≥3 COVID-19 vaccine doses.

**Implications:** In a setting with high levels of COVID-19 vaccine and booster uptake, receipt of nirmatrelvir-ritonavir 0–5 days after symptom onset was associated with substantial reductions in risk of hospital admission or death within 30 days after a positive outpatient SARS-CoV-2 test.

**Funding:** US Centers for Disease Control and Prevention, US National Institutes of Health

## INTRODUCTION

In conjunction with vaccination as a primary prevention strategy, therapeutic drugs to prevent severe outcomes of SARS-CoV-2 infection are of key importance to current efforts aimed at mitigating the burden of COVID-19. In early trials, neutralizing monoclonal antibody therapies^1–3^ and remdesivir,^4^ an antiviral agent repurposed for treatment of COVID-19 cases, showed success in preventing hospital admissions when administered early in patients’ clinical course. However, the need to deliver these treatments by intravenous infusion has limited their utility for broad implementation in ambulatory care settings. Additionally, many monoclonal antibody therapies have been rendered ineffective by continuing mutations in the Spike protein of SARS-CoV-2.^5^ In the randomized phase II/III EPIC-HR trial among high-risk, unvaccinated, and non-hospitalized adults,^6^ oral nirmatrelvir-ritonavir (Paxlovid™, Pfizer, Inc.) reduced the risk of COVID-19 related hospital admission by 89% over 28 days following treatment dispense. Based on this evidence, the US Food and Drug Administration issued Emergency Use Authorization for nirmatrelvir-ritonavir for adults and children aged ≥12 years weighing ≥40 kilograms who are at high risk for progression to severe COVID-19.

Amid the broad implementation of nirmatrelvir-ritonavir as a treatment option for COVID-19 cases diagnosed in ambulatory settings,^7^ evidence is needed on effectiveness in preventing severe disease under real-world conditions of use. In recent observational studies, nirmatrelvir-ritonavir was estimated to confer 21–79% reductions in risk of hospital admission or other severe endpoints, with the strongest evidence of clinical benefit among high-risk subgroups including older adult cases, obese cases, and cases not known to have immunity to SARS-CoV-2 from prior infection or vaccination.^8–14^ Factors explaining this wide range of effectiveness estimates remain uncertain. Whereas timing of administration may be of crucial importance to the effectiveness of antiviral therapies, prior effectiveness studies have lacked symptoms data, making it difficult to compare results across studies potentially enrolling cases with heterogeneous clinical status. Expansions in nirmatrelvir-ritonavir access and availability have led to widening uptake among a broader patient population including non-elderly and vaccinated cases, as well as those without high-risk chronic comorbid conditions, further contributing to potential variation across studies.^15,16^ Last, rollout of nirmatrelvir-ritonavir has occurred during periods dominated by circulation of the BA.2, BA.4, and BA.5 SARS-CoV-2 Omicron lineages, when most individuals in the United States and other countries have shown antibody evidence of prior SARS-CoV-2 infection.^17^ These circumstances present a notable contrast to the EPIC-HR trial, which (1) randomized participants to treatment or placebo within the first 5 days after symptom onset; (2) enrolled only unvaccinated high-risk (e.g. older, chronically ill, or obese) adults without a history of prior infection; and (3) was conducted while the Delta (B.1.617.2) variant, which has been associated with greater disease severity,^18,19^ remained the predominant circulating SARS-CoV-2 lineage.

While data from the United States have shown that COVID-19 related emergency department (ED) presentations and hospital admissions are infrequent among cases receiving nirmatrelvir-ritonavir in ambulatory care settings,^18,20^ systematic assessments of effectiveness remain lacking. We therefore aimed to measure the effectiveness of nirmatrelvir-ritonavir in preventing severe outcomes of SARS-CoV-2 infection among cases ascertained via outpatient testing within a large, integrated US healthcare system.

## METHODS

### Setting and eligibility criteria

Our study was conducted within Kaiser Permanente Southern California (KPSC), a comprehensive healthcare system providing integrated care across virtual, outpatient, emergency department (ED), and inpatient settings to 4.7 million members (∼19% of the population of southern California). Members of KPSC are enrolled through employer-provided, pre-paid, and federally-sponsored insurance plans and represent the diverse socioeconomic characteristics and racial/ethnic backgrounds of the geographic areas served.^21,22^ Electronic health records (EHRs) capture all within-network care delivery including diagnoses, pharmacy dispenses, laboratory tests and results, and vaccinations received. Care received out of network is captured through insurance claim reimbursements. COVID-19 vaccinations received outside KPSC were captured through linkage with the California Immunization Registry, (to which providers were required to report all COVID-19 vaccine administrations within 24 hours^23^) as well as other health systems using the Epic EHR system.

Although Emergency Use Authorization for nirmatrelvir-ritonavir was granted in December, 2021, real-world uptake remained low through the first months of 2022 due to limited supplies, and treatment was initially prioritized to KPSC patients who were immunocompromised or on immunosuppressive medications, aged ≥65 years and incompletely vaccinated, or aged ≥65 years with risk factors for severe disease. By April, 2022, nirmatrelvir-ritonavir was available to KPSC patients receiving a positive SARS-CoV-2 test result in any outpatient setting or reporting a positive at-home antigen test, with no requirement for an in-person appointment. Patients taking statins or hormonal contraceptives containing ethinyl estradiol were instructed to withhold taking these medications while taking nirmatrelvir-ritonavir, while those taking medication for HIV infection were instructed to continue taking HIV antiretroviral medications and arrange follow-up with care providers to monitor for side effects. Patients taking other concomitant medications with potential interactions were referred for clinical evaluation to determine appropriate therapeutic options, including withholding of other medications, dose adjustment, or increased monitoring to assess drug interactions.

Cases were eligible for inclusion in the current study if they: (1) received a positive SARS-CoV-2 polymerase chain reaction (PCR) test result, defined as their index test, between 8 April and 7 October, 2022 (a period when ≥5% of outpatient-diagnosed cases at KPSC received nirmatrelvir-ritonavir each week); (2) had no prior positive test result within the preceding 90 days; were ≥12 years old at the time of their index test; (3) were not hospitalized at the time of their index test or within the preceding 7 days; and (4) had ≥1 year of continuous enrollment in KPSC health plans before their index test (allowing for a 45-day gap during previous membership due to potential delays in renewal). We included follow-up for clinical endpoints in this study through 30 or 60 days after cases’ index test (as detailed below), through disenrollment, or through 20 October, 2022, whichever occurred earliest.

### Exposures

The primary exposures were receipt of nirmatrelvir-ritonavir 0–5 days after symptom onset, and receipt of nirmatrelvir-ritonavir at any time after testing positive for SARS-CoV-2 (regardless of the presence or timing of symptoms). We considered participants to be exposed to nirmatrelvir-ritonavir beginning on the date of treatment dispense, as recorded in KPSC pharmacy records or adjudicated out-of-network insurance claims. Cases who received nirmatrelvir-ritonavir ≥1 day after their index test were considered unexposed during the time between their index test and date of treatment dispense.

Data on symptoms associated with SARS-CoV-2 infection were abstracted from structured questionnaires administered with each SARS-CoV-2 test order, and from unstructured text fields within EHRs, as described previously.^24^ The date of SARS-CoV-2 symptom onset was defined as the earliest date that cases reported acute fever, cough, headache, fatigue, dyspnea, chills, sore throat, myalgia, anosmia, diarrhea, vomiting/nausea, or abdominal pain within 14 days before or after their index test date. If new-onset symptoms were not recorded within this period, we categorized cases as not experiencing acute COVID-19 symptoms in association with their infection. To support exploratory analyses addressing effectiveness of nirmatrelvir-ritonavir at differing clinical stages in cases’ clinical course, we defined additional subgroups including cases treated 0–3 days after symptom onset (the primary exposure assessed in the EPIC-HR trial^6^); cases treated ≥6 days after symptom onset or in the absence of acute COVID-19 symptoms; and cases treated at any time after symptom onset. As an additional exploratory analysis aiming to emulate the design of the EPIC-HR trial, in which cases were randomized to treatment or placebo at the point of testing, we distinguished courses dispensed on the day of testing, censoring observations from cases who received treatment at later points in time.

Additional variables recorded at the time of participants’ index test included: age, sex, race/ethnicity (non-Hispanic White, Black, Asian, or Pacific Islander; Hispanic [belonging to any race]; or other/unknown/multiple race), body mass index (BMI), current or former cigarette smoking, socioeconomic status (measured by census-tract neighborhood deprivation index^25^ quintiles), COVID-19 vaccine doses received, prior documented SARS-CoV-2 infection, comorbid conditions (from which we computed Charlson comorbidity index values for each case as a summary measure of risk^26–28^), healthcare utilization in the prior year (comprising interactions across outpatient, emergency department [ED], and inpatient settings), and receipt of other vaccines (including 2021-22 season influenza vaccination, pneumococcal polysaccharide vaccine, and pneumococcal conjugate vaccine; **Table 1**).

**Table 1:**
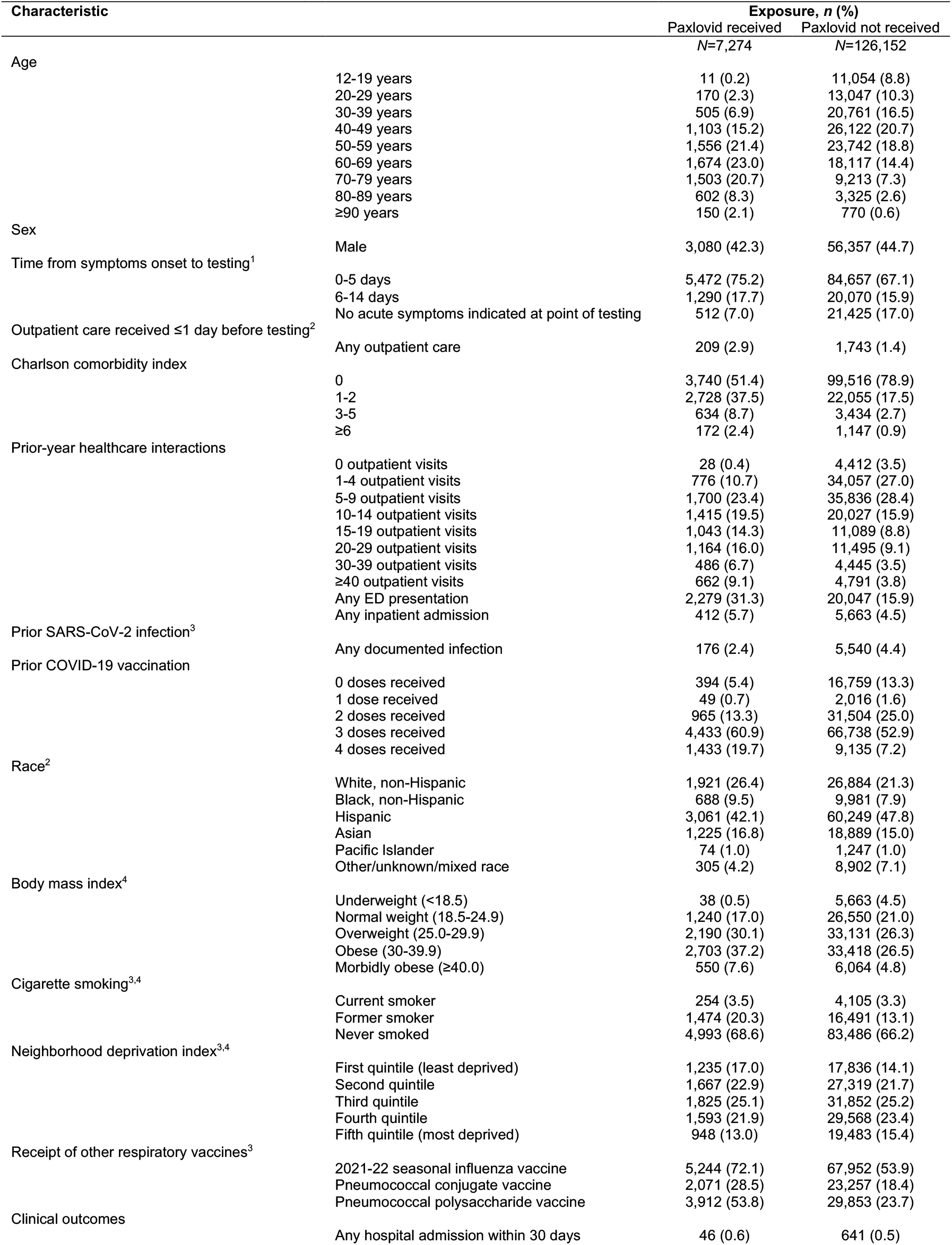
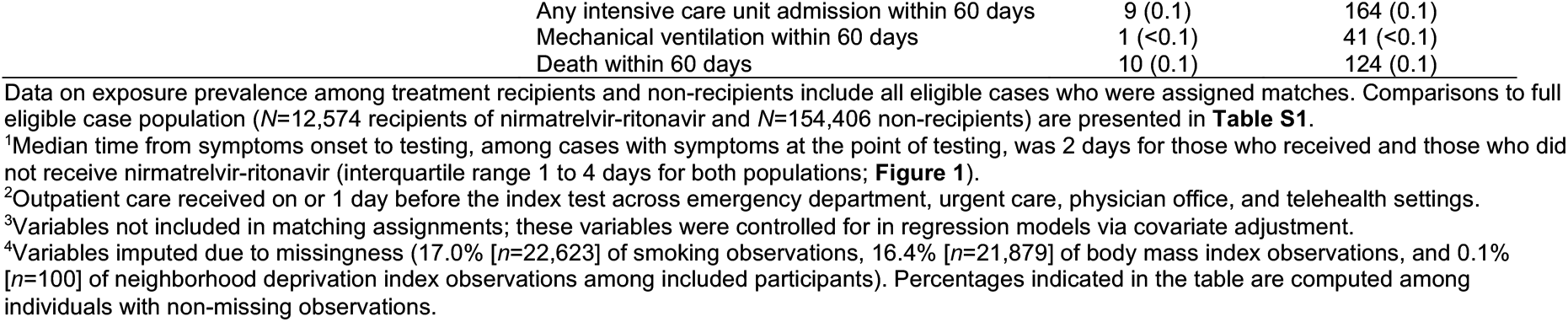
Characteristics of the analytic sample.

| Characteristic |  | Exposure, n (%) |  |
| --- | --- | --- | --- |
|  |  | Paxlovid received<br>N=7,274 | Paxlovid not received<br>N=126,152 |
| Age |  |  |  |
|  | 12-19 years | 11 (0.2) | 11,054 (8.8) |
|  | 20-29 years | 170 (2.3) | 13,047 (10.3) |
|  | 30-39 years | 505 (6.9) | 20,761 (16.5) |
|  | 40-49 years | 1,103 (15.2) | 26,122 (20.7) |
|  | 50-59 years | 1,556 (21.4) | 23,742 (18.8) |
|  | 60-69 years | 1,674 (23.0) | 18,117 (14.4) |
|  | 70-79 years | 1,503 (20.7) | 9,213 (7.3) |
|  | 80-89 years | 602 (8.3) | 3,325 (2.6) |
|  | ≥90 years | 150 (2.1) | 770 (0.6) |
| Sex | Male | 3,080 (42.3) | 56,357 (44.7) |
| Time from symptoms onset to testing<sup>1</sup> | 0-5 days | 5,472 (75.2) | 84,657 (67.1) |
|  | 6-14 days | 1,290 (17.7) | 20,070 (15.9) |
|  | No acute symptoms indicated at point of testing | 512 (7.0) | 21,425 (17.0) |
| Outpatient care received ≤1 day before testing<sup>2</sup> | Any outpatient care | 209 (2.9) | 1,743 (1.4) |
| Charlson comorbidity index | 0 | 3,740 (51.4) | 99,516 (78.9) |
|  | 1-2 | 2,728 (37.5) | 22,055 (17.5) |
|  | 3-5 | 634 (8.7) | 3,434 (2.7) |
|  | ≥6 | 172 (2.4) | 1,147 (0.9) |
| Prior-year healthcare interactions | 0 outpatient visits | 28 (0.4) | 4,412 (3.5) |
|  | 1-4 outpatient visits | 776 (10.7) | 34,057 (27.0) |
|  | 5-9 outpatient visits | 1,700 (23.4) | 35,836 (28.4) |
|  | 10-14 outpatient visits | 1,415 (19.5) | 20,027 (15.9) |
|  | 15-19 outpatient visits | 1,043 (14.3) | 11,089 (8.8) |
|  | 20-29 outpatient visits | 1,164 (16.0) | 11,495 (9.1) |
|  | 30-39 outpatient visits | 486 (6.7) | 4,445 (3.5) |
|  | ≥40 outpatient visits | 662 (9.1) | 4,791 (3.8) |
|  | Any ED presentation | 2,279 (31.3) | 20,047 (15.9) |
|  | Any inpatient admission | 412 (5.7) | 5,663 (4.5) |
| Prior SARS-CoV-2 infection<sup>3</sup> | Any documented infection | 176 (2.4) | 5,540 (4.4) |
| Prior COVID-19 vaccination | 0 doses received | 394 (5.4) | 16,759 (13.3) |
|  | 1 dose received | 49 (0.7) | 2,016 (1.6) |
|  | 2 doses received | 965 (13.3) | 31,504 (25.0) |
|  | 3 doses received | 4,433 (60.9) | 66,738 (52.9) |
|  | 4 doses received | 1,433 (19.7) | 9,135 (7.2) |
| Race<sup>2</sup> | White, non-Hispanic | 1,921 (26.4) | 26,884 (21.3) |
|  | Black, non-Hispanic | 688 (9.5) | 9,981 (7.9) |
|  | Hispanic | 3,061 (42.1) | 60,249 (47.8) |
|  | Asian | 1,225 (16.8) | 18,889 (15.0) |
|  | Pacific Islander | 74 (1.0) | 1,247 (1.0) |
|  | Other/unknown/mixed race | 305 (4.2) | 8,902 (7.1) |
| Body mass index<sup>4</sup> | Underweight (<18.5) | 38 (0.5) | 5,663 (4.5) |
|  | Normal weight (18.5-24.9) | 1,240 (17.0) | 26,550 (21.0) |
|  | Overweight (25.0-29.9) | 2,190 (30.1) | 33,131 (26.3) |
|  | Obese (30-39.9) | 2,703 (37.2) | 33,418 (26.5) |
|  | Morbidly obese (≥40.0) | 550 (7.6) | 6,064 (4.8) |
| Cigarette smoking<sup>3,4</sup> | Current smoker | 254 (3.5) | 4,105 (3.3) |
|  | Former smoker | 1,474 (20.3) | 16,491 (13.1) |
|  | Never smoked | 4,993 (68.6) | 83,486 (66.2) |
| Neighborhood deprivation index<sup>3,4</sup> | First quintile (least deprived) | 1,235 (17.0) | 17,836 (14.1) |
|  | Second quintile | 1,667 (22.9) | 27,319 (21.7) |
|  | Third quintile | 1,825 (25.1) | 31,852 (25.2) |
|  | Fourth quintile | 1,593 (21.9) | 29,568 (23.4) |
|  | Fifth quintile (most deprived) | 948 (13.0) | 19,483 (15.4) |
| Receipt of other respiratory vaccines<sup>3</sup> | 2021-22 seasonal influenza vaccine | 5,244 (72.1) | 67,952 (53.9) |
|  | Pneumococcal conjugate vaccine | 2,071 (28.5) | 23,257 (18.4) |
|  | Pneumococcal polysaccharide vaccine | 3,912 (53.8) | 29,853 (23.7) |
| Clinical outcomes | Any hospital admission within 30 days | 46 (0.6) | 641 (0.5) |

|  |  |  |
| --- | --- | --- |
| Any intensive care unit admission within 60 days | 9 (0.1) | 164 (0.1) |
| Mechanical ventilation within 60 days | 1 (<0.1) | 41 (<0.1) |
| Death within 60 days | 10 (0.1) | 124 (0.1) |

<sup>1</sup>Median time from symptoms onset to testing, among cases with symptoms at the point of testing, was 2 days for those who received and those who did not receive nirmatrelvir-ritonavir (interquartile range 1 to 4 days for both populations; **Figure 1**).
<sup>2</sup>Outpatient care received on or 1 day before the index test across emergency department, urgent care, physician office, and telehealth settings.
<sup>3</sup>Variables not included in matching assignments; these variables were controlled for in regression models via covariate adjustment.
<sup>4</sup>Variables imputed due to missingness (17.0% [ $n=22,623$ ] of smoking observations, 16.4% [ $n=21,879$ ] of body mass index observations, and 0.1% [ $n=100$ ] of neighborhood deprivation index observations among included participants). Percentages indicated in the table are computed among individuals with non-missing observations.

### Endpoints

The primary endpoint of this study was hospital admission or death due to any cause within 30 days after participants’ index positive SARS-CoV-2 test. This endpoint was selected to maximize comparability to the endpoint of the EPIC-HR trial,^6^ which monitored participants for COVID-19 related hospital admission or death due to any cause after a positive test. Throughout the study period, KPSC implemented a home-based monitoring program for COVID-19 cases aiming to preserve hospital capacity by standardizing admission criteria, as described previously.^29^ Briefly, cases judged to be at high risk of severe outcomes based on their risk factors and clinical status at the point of testing were given medical-grade pulse oximeters and thermometers, and monitored for clinical deterioration on the basis of daily readings, reported through either a mobile application or daily telephone calls from providers. Standardized criteria were used for subsequent ED referral and inpatient admission, including SpO_2_ readings, respiratory rate, and patient-reported shortness of breath or other clinical signs or symptoms. Hospital admission was thus considered to represent an internally-consistent disease severity threshold within our sample monitored from the point of outpatient testing.

As a secondary endpoint indicating progression to more severe disease, we also evaluated intensive care unit (ICU) admission, mechanical ventilation, or death within 60 days after participants’ index test date. We selected a 60-day interval for evaluation of this secondary endpoint to accommodate potential lags in the occurrence of severe clinical outcomes.^21^

### Design and statistical analysis

We compared clinical outcomes associated with nirmatrelvir-ritonavir receipt in a matched cohort framework, monitoring participants from the point of their index test to the occurrence of each study endpoint or censoring (due end of follow-up, study end, or disenrollment, whichever occurred earlier). We updated participants’ treatment assignments at the date of treatment dispense if this occurred ≥1 day after the index test. For each study endpoint, we defined treatment effectiveness as (1 − adjusted hazards ratio [aHR]) × 100%, for the aHR comparing outcomes among cases who had received or had not received nirmatrelvir-ritonavir. Effectiveness estimates based on the aHR throughout cases’ full follow-up duration should be interpreted as weighted averages of the instantaneous aHRs comparing cases who had received or had not received treatment at each point in time.^30,31^ We estimated the aHR and accompanying 95% confidence interval (CI) via Cox proportional hazards models, using the Andersen-Gill extension^32^ to update time-varying exposures. We estimated cluster-robust standard errors to account for multiple observations from cases whose treatment status changed during follow-up. We verified the proportional hazards assumption by testing for non-zero slopes of Schoenfeld residuals from fitted models.^33^

To mitigate confounding driven by factors associated with cases’ likelihood of both receiving nirmatrelvir-ritonavir and experiencing severe clinical outcomes, we constructed a directed acyclic graph identifying a minimal set of covariates for statistical adjustment (**Figure S1**). We defined regression strata (matches) among cases on the basis of their week of SARS-CoV-2 testing; age (within bands of 12-19, 20-29, 30-39, …, 80-89, and ≥90 years); sex; clinical status; healthcare utilization in the year prior to the index test (0-4, 5-14, 15-29, or ≥30 outpatient visits and ≥1 ED presentation or inpatient admission); COVID-19 vaccine doses received; presence of comorbidities (Charlson comorbidity index values of 0, 1–2, 3–5, or ≥6) and body mass index category (underweight, normal weight, overweight, obese, or morbidly obese, as defined in **Table 1**). We measured clinical status according to two criteria: (1) receipt of any clinical care in association with testing (across ED, urgent care, outpatient, or telehealth settings), and (2) days from symptom onset, or absence of acute symptoms, at the start of the observation interval. As prescribing guidelines have assigned differing priority to cases with one or more risk factors (e.g., age, obesity, comorbidity, or lack of vaccination),^34^ and interactions are present in the effects of these risk factors on cases’ likelihood of severe disease,^35,36^ this approach was selected to allow differing baseline hazards across all combinations of the listed covariates. We further controlled for race/ethnicity, current or former cigarette smoking, documentation of prior SARS-CoV-2 infection, neighborhood deprivation index quintile, and receipt of other vaccines (as an additional measure of healthcare-seeking behavior) as covariates in regression models, as effect modification was considered less likely among these variables. We repeated analyses in subgroups who received ≥2 or ≥3 COVID-19 vaccine doses.

Because smoking status, BMI, and census-tract neighborhood deprivation index measures may be missing within KSPC records,^19^ we conducted multiple imputation to sample from the joint distribution of these variables with respect to all other observed characteristics of cases.^37^ We repeated analyses across each of 5 independent pseudo-datasets completed via imputation of missing observations. Reported results pool parameter estimates across analyses undertaken with each of the 5 pseudo-datasets according to Rubin’s rules.^38^

All analyses were performed using R software (version 4.2.1; R Foundation for Statistical Computing, Vienna, Austria). The study was approved by the KPSC Institutional Review Board, which granted a waiver of informed consent.

## RESULTS

In total, 197,484 KPSC members without a prior positive test result within 90 days received positive SARS-CoV-2 molecular test results during the study period, among whom 166,980 were aged ≥12 years with ≥1 year of continuous enrollment before their test date and thus eligible for inclusion (**Table S1**). Within this population, 12,574 (7.5% of 166,980) cases received nirmatrelvir-ritonavir at any point in their clinical course, among whom 10,038 (79.8% of 12,574) were tested within 0–5 days after symptom onset, 1,755 (14.0%) were tested ≥6 days after symptom onset, and 781 (6.2%) did not have acute SARS-CoV-2 symptoms recorded at the point of testing (**Figure 1**). Among all treated cases, 11,653 (92.7% of 12,574) did not have clinical care interactions with providers in ED, urgent care, outpatient, or telehealth settings in association with their index test. Among 154,406 cases who did not receive nirmatrelvir-ritonavir, 101,537 (65.8%) were tested within 0–5 days of symptom onset, 26,903 (17.4%) were tested ≥6 days after symptom onset, 25,966 (16.8%) did not have acute symptoms recorded at the point of testing, and 146,609 (95.0%) did not have interactions with outpatient care providers in association with testing (**Table S1**). Recipients of nirmatrelvir-ritonavir were generally older than non-recipients (60.3% [7,582] of recipients and 28.8% [44,500] of non-recipients were ≥60 years old), were more likely to have chronic comorbid conditions (57.6% [7,237] of recipients and 28.1% [43,314] of non-recipients; **Table S2**), were more likely to be obese or morbidly obese (44.6% [5,604] of recipients and 32.7% [50,544] of non-recipients), had higher levels of healthcare utilization within the prior year (40.7% [5,115] of recipients and 24.2% [37,407] of non-recipients had ≥1 ED presentation or inpatient admission), and were more likely to have received COVID-19 vaccines (90.8% [11,411] of recipients and 83.0% [128,090] of non-recipients had received ≥2 doses, while 7.4% [932] of recipients and 14.5% [22,338] of non-recipients had received 0 doses). Differences between recipients and non-recipients in race/ethnicity, cigarette smoking, and community socioeconomic characteristics were less clearly pronounced. Among cases aged 12-39 years or 40-64 years, presence of comorbidities and high BMI were associated with greater increases in the likelihood of receiving nirmatrelvir-ritonavir than among cases aged ≥65 years (**Table S3**).

**Figure 1:**
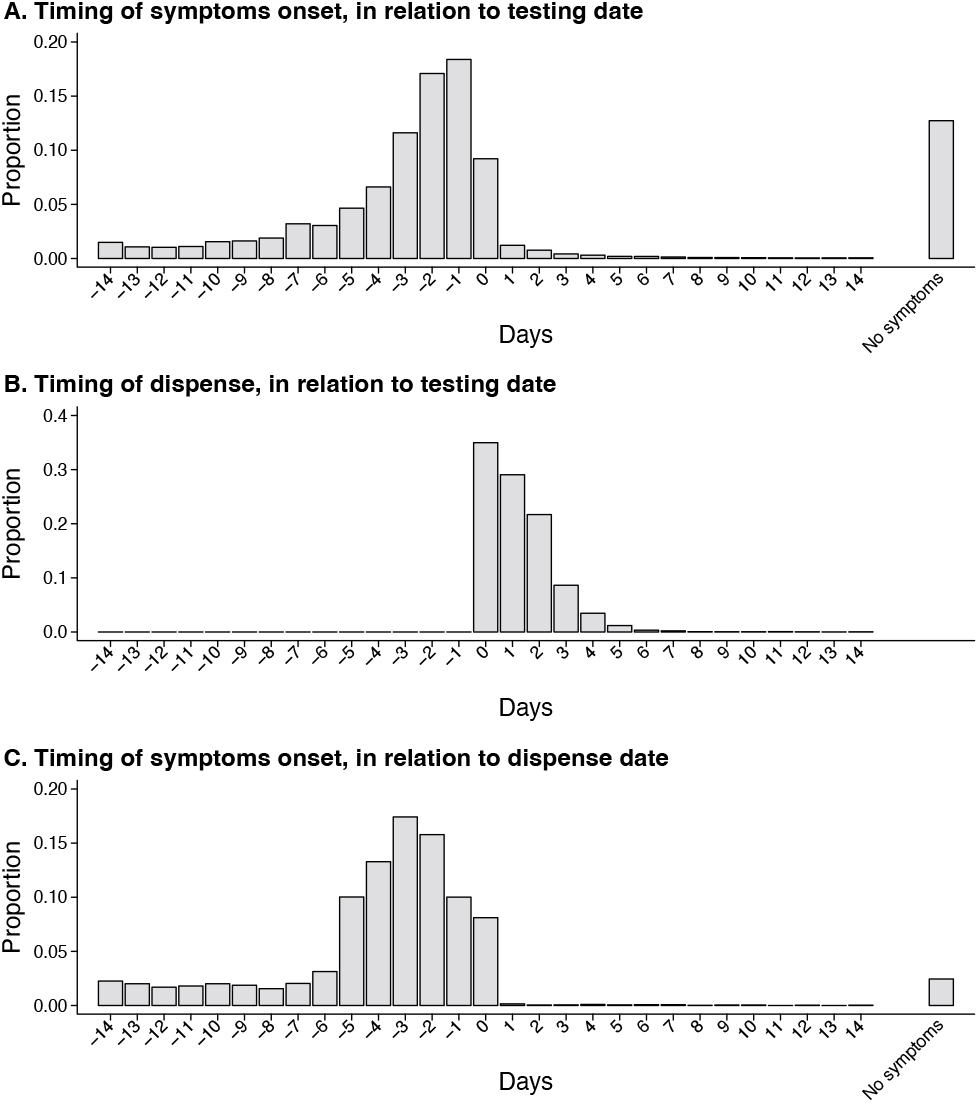
Timing of SARS-CoV-2 testing and nirmatrelvir-ritonavir dispense. We present distributions of: (**A**) times from symptom onset to cases’ index positive test for SARS-CoV-2 (defined as day 0), for all cases; (**B**) times from testing to nirmatrelvir-ritonavir dispense (defined as day 0), for cases who received treatment; and (**C**) times from symptom onset to nirmatrelvir-ritonavir dispense (defined as day 0), for cases who received treatment.

The proportion of cases receiving nirmatrelvir-ritonavir increased with time throughout the study period. Whereas the weeks from 31 December, 2021 to 7 April, 2022 saw dramatic increases in nirmatrelvir-ritonavir dispenses first among cases aged ≥65 years, those with comorbid conditions, and those with BMI values indicating obesity or morbid obesity, differences in the relative likelihood of nirmatrelvir-ritonavir dispense among these groups were became pronounced for cases diagnosed between 8 April to 7 October, 2022 (**Figure 2**).

**Figure 2:**
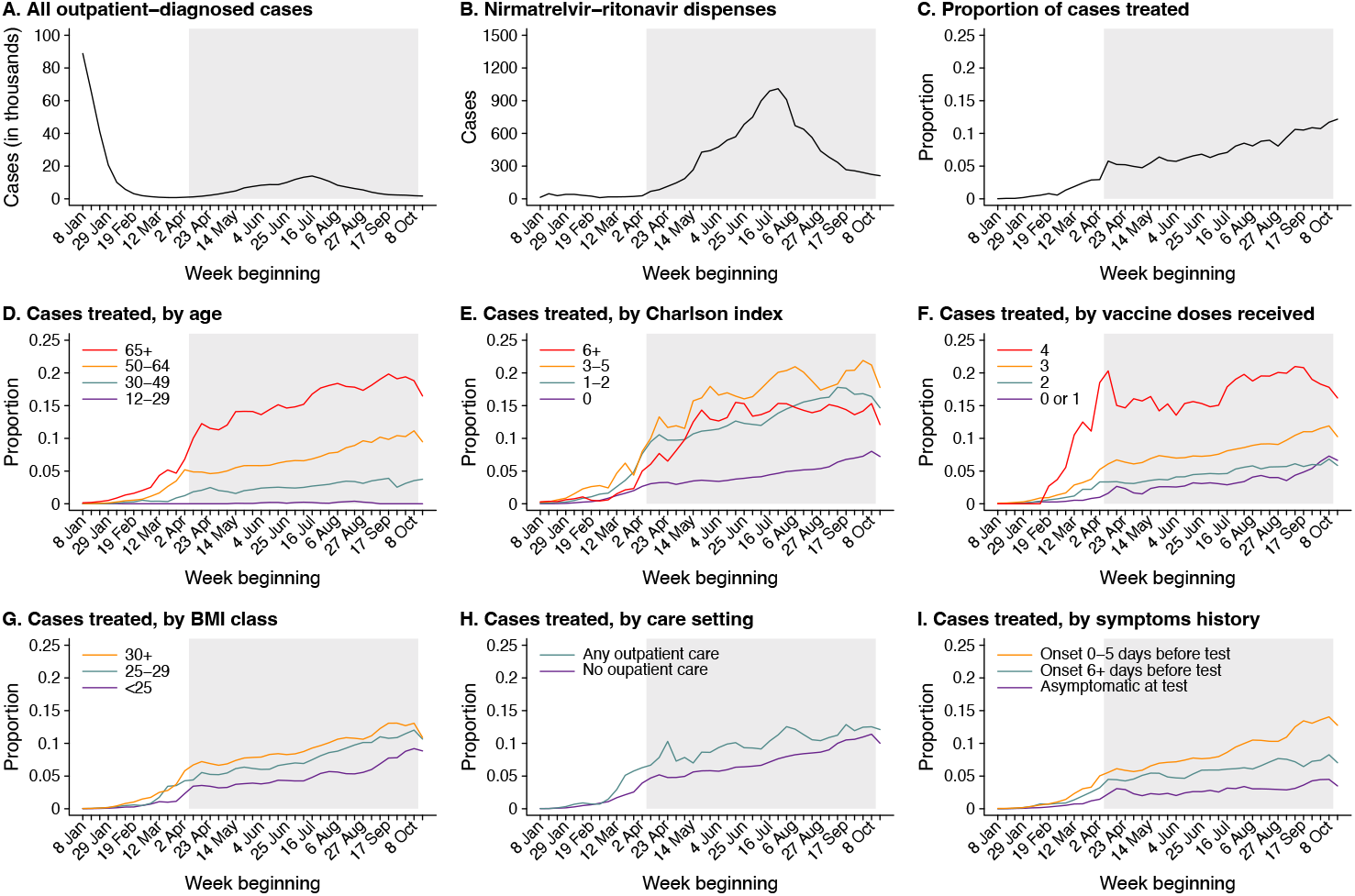
Receipt of nirmatrelvir-ritonavir among cases over time and according to risk factors. Indexing cases by week of their index positive test, panels in the top row illustrate (**A**) total new-onset positive SARS-CoV-2 cases identified via polymerase chain reaction testing in outpatient settings; (**B**) the number of outpatient-identified cases receiving nirmatrelvir-ritonavir; and (**C**) the proportion of outpatient-identified cases receiving oral nirmatrelvir-ritonavir. Panels in the middle and bottom rows disaggregate the proportion of outpatient-identified cases receiving oral nirmatrelvir-ritonavir according to (**D**) age group; (**E**) Charlson comorbidity index values (for which we enumerate specific underlying comorbid conditions within the sample in **Table S2**); (**F**) number of vaccine doses received; (**G**) body mass index; (**H**) receipt of outpatient clinical care in association with testing, defined as any provider interaction in an ED, urgent care, outpatient/physician office, or telehealth setting; and (**I**) presence and timing of potential COVID-19 symptoms at the point of testing. Grey shaded areas delineate testing dates for which cases were included in the study period. Lines in panels **D-I** present 3-week moving averages incorporating data from 7 days before and after each study week.

Among all eligible cases, 7,274 (57.8% of 12,574) nirmatrelvir-ritonavir recipients and 126,152 (81.7% of 154,406) non-recipients were retained in analyses with ≥1 eligible match based on their week of SARS-CoV-2 testing, age, sex, clinical status, prior-year healthcare utilization, COVID-19 vaccine doses received, Charlson comorbidity index, and BMI (**Table 1**; **Table S4**). In comparison to the full eligible population, cases for whom matches were identified were less likely to have received outpatient care in association with testing (1.5% [1,952] versus 5.2% [8,718]), were slightly younger (26.5% [35,354] versus 31.2% [52,082] aged ≥60 years), were less likely to have comorbid conditions (22.6% [30,170] versus 69.7% [50,551]), and were less likely to have had ≥1 ED presentation or inpatient admission in the prior year (18.9% [25,272] versus 25.5% [42,522]). In total, the frequency of hospital admission, ICU admission, mechanical ventilation, and death during follow-up was 0.5% (687), 0.1% (173), 42 (<0.1%), and 134 (0.1%), respectively, within the population retained in analyses, as compared to 0.9% (1,440), 0.3% (429), 0.1% (133), and 423 (0.3%), respectively, within the full eligible population. Differences between recipients and non-recipients of nirmatrelvir-ritonavir within the analytic sample resembled those within the full eligible population (**Table 1, Table S1, Table S2, Table S4**).

The primary endpoint of hospital admission or death due to any cause within 30 days from the positive index test occurred among 51 (0.7% of 7,274) nimatrelvir-ritonavir recipients and 695 (0.5% of 126,152) non-recipients (**Table S5**). Disenrollment or censoring before 30 days, or before occurrence of the primary endpoint, occurred for 212 (2.9% of 7,274) treated participants and 2,790 (2.1% of 126,152) participants who did not receive treatment. After adjustment for the aforementioned differences in risk status among treated and untreated cases, receipt of nirmatrelvir-ritonavir 0–5 days after symptom onset was associated with 79.6% (33.9% to 93.8%) effectiveness against progression to the primary endpoint of hospital admission or death due to any cause within 30 days after cases’ index test (**Table 2**). Courses administered at any time, regardless of the presence or timing of symptoms, were associated with 53.6% (6.6% to 77.0%) effectiveness against progression to the same endpoint. Among cases who received nirmatrelvir-ritonavir, cases who progressed to hospital admission or death within 30 days of their index test were older (82.4% [42/51] versus 53.8% [3,887/7,223] treated cases experiencing and not experiencing the outcome, respectively); more likely to have been tested ≥6 days after symptom onset (41.2% [21] versus 17.6% [1,269] of treated cases experiencing or not experiencing the outcome, respectively); and more likely to have comorbid conditions (82.4% [42] versus 48.3% [3,492] of treated cases experiencing or not experiencing the outcome; **Table S6**).

**Table 2:** Effectiveness of nirmatrelvir-ritonavir in preventing progression to severe disease endpoints.

| Endpoint | Timing of dispense in relation to symptoms onset<sup>1</sup> | Discordant pairs<sup>2</sup> |  | Effectiveness |  |
| --- | --- | --- | --- | --- | --- |
|  |  | Outcome observed for recipient, non-recipient censored | Outcome observed for non-recipient, recipient censored | Estimate, % (95% confidence interval) | p-value (two-sided) |
| All-cause hospital admission or death within 30 days from positive SARS-CoV-2 test | 0-5 days after symptoms onset | 8 | 11 | 79.6 (33.9, 93.8) | <0.01 |
|  | At any time, regardless of symptoms | 26 | 23 | 53.6 (6.6, 77.0) | 0.03 |
| All-cause intensive care unit admission, mechanical ventilation, or death within 60 days from positive SARS-CoV-2 test | 0-5 days after symptoms onset | 2 | 7 | 89.2 (–25.0, 99.3) | 0.07 |
|  | At any time, regardless of symptoms | 10 | 11 | 84.1 (18.8, 96.9) | 0.03 |

<sup>1</sup>We present effectiveness estimates for alternative subgroups, defined by time from symptoms onset to testing and presence or absence of symptoms, in **Table S8** and **Table S9**.
<sup>2</sup>Reasons for censoring and lengths of follow-up are enumerated in **Table S5**.

For the secondary endpoint of ICU admission, mechanical ventilation, or death within 60 days after cases’ index test date, receipt of nirmatrelvir-ritonavir was associated with 89.2% (–25.0% to 99.3%) effectiveness when administered within 0–5 days of symptom onset (**Table 2**). For dispenses occurring at any time in the clinical course, effectiveness against the same endpoint was 84.1% (18.8% to 96.9%). Attributes distinguishing cases who experienced this secondary endpoint from those who did not resembled attributes predicting hospital admission or death within 30 days after cases’ index test (**Table S7**).

Exploratory analyses identified varying estimates of treatment effectiveness according to the timing of dispense in relation to symptom onset. Nirmatrelvir-ritonavir was associated with 89.6% (50.2% to 97.8%) effectiveness against progression to the primary endpoint of hospital admission or death due to any cause within 30 days after a positive test for cases who received treatment on the day of their index test and 0–5 days from symptom onset, consistent with the design of the EPIC-HR trial (**Table S8**). For all cases dispensed treatment on the day of their index test, effectiveness against the same endpoint was 77.7% (31.3% to 92.7%). Treatment dispenses occurring 0–3 days after symptom onset and at any time after symptom onset were associated with 81.1% (34.1% to 94.6%) and 59.5% (12.8% to 81.2%) effectiveness against hospital admission or death within 30 days after the index test, respectively. Effectiveness of treatment dispenses occurring ≥6 days after symptom onset or in the absence of acute symptoms was 43.8% (–33.3% to 81.7%; **Table S9**).

Cases tested and treated at later stages of their clinical course were generally older than those tested or treated within 0– 5 days, more likely to receive outpatient care in association with testing, and more likely to have chronic comorbid conditions (**Table S10, Table S11**, and **Table S12**).

In subgroup analyses of cases who had received ≥2 and ≥3 COVID-19 vaccine doses, effectiveness of nirmatrelvir-ritonavir against the primary endpoint of hospital admission or death within 30 days was 83.1% (30.4% to 95.9%) and 92.2% (52.0% to 98.7%), respectively, when treatment dispense occurred 0–5 days from symptom onset (**Table 3**). Effectiveness against the same endpoint for treatment dispenses occurring at any time was 55.3% (6.6% to 78.7%) for cases who had received ≥2 COVID-19 vaccine doses, and 66.5% (24.0% to 85.3%) for cases who had received ≥3 COVID-19 vaccine doses.

**Table 3:**
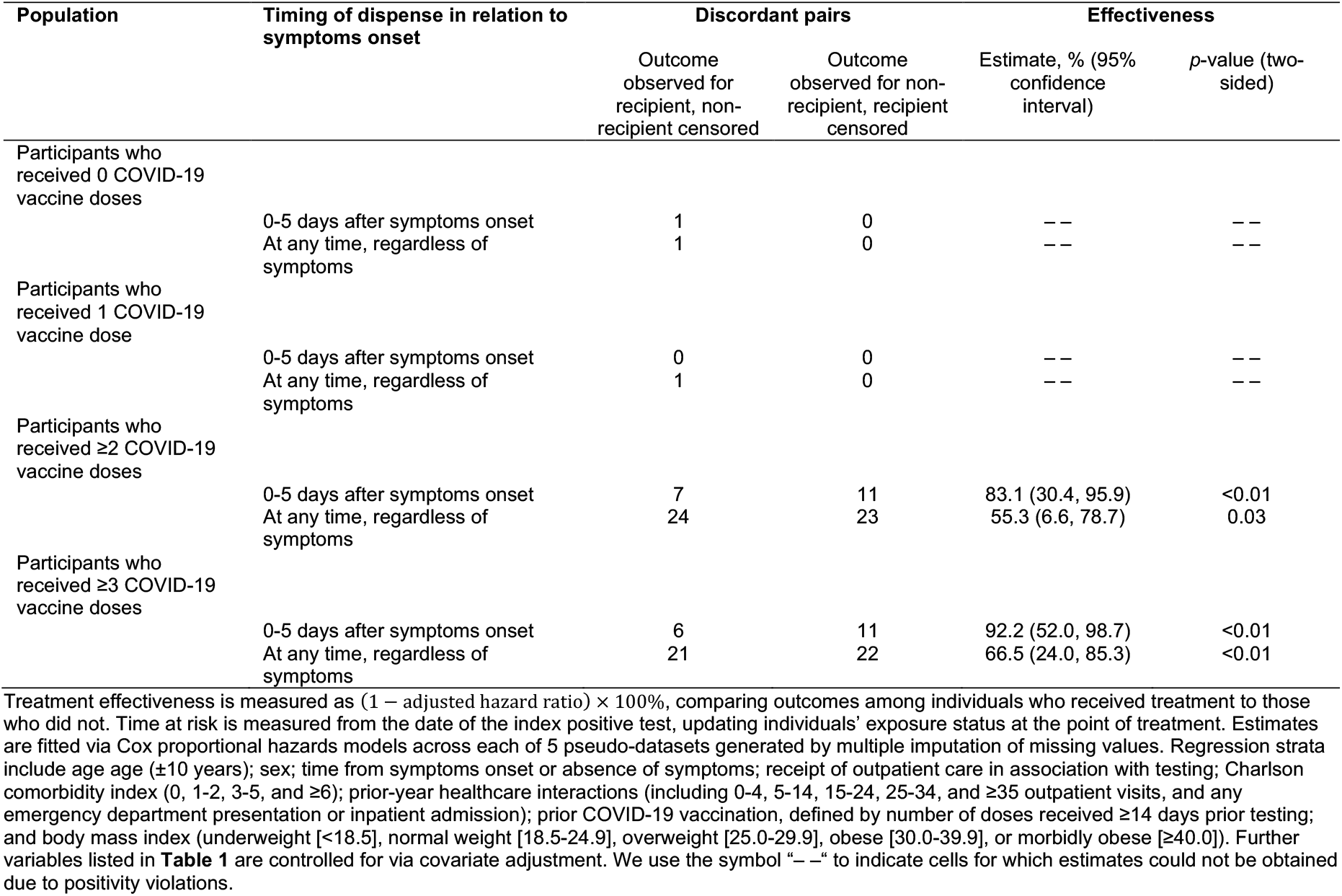
Effectiveness of Paxlovid in preventing progression to hospital admission or death within 30 days from positive SARS-CoV-2 test, according to prior COVID-19 vaccination or documented history of SARS-CoV-2 infection.

## DISCUSSION

In this cohort study conducted among a highly vaccinated population in a large US healthcare system, nirmatrelvir-ritonavir was effective in preventing hospitalization or death within 30 days following a positive outpatient test for SARS-CoV-2 infection. For treatment courses administered at any point during the clinical course, including in the absence of symptoms, effectiveness against the primary endpoint of hospital admission or death due to any cause was 53.6% (6.6% to 77.0%). Early treatment was associated with the greatest clinical benefit: dispenses occurring within 0–5 and 0–3 days from symptom onset were associated with 79.6% (33.9% to 93.8%) and 81.1% (34.1% to 94.6%) effectiveness against this endpoint, respectively. Among cases tested 0–5 days following symptom onset, receipt of nirmatrelvir-ritonavir on the day of testing was associated with 89.6% (50.2% to 97.8%) effectiveness against the primary endpoint. This finding closely emulates results of the EPIC-HR trial, in which cases who were randomized to treatment or placebo on the day of their test (and 0–5 days after symptom onset) experienced an 87.8% reduction in risk of COVID-19 associated hospital admission or death due to any cause.^6^ In our study population, cases treated ≥6 days after symptom onset constituted only 22.2% of nirmatrelvir-ritonavir recipients, but accounted for 45.7% of recipients who were hospitalized, 70.0% of recipients who required intensive care or mechanical ventilation, and 60.0% of recipients who died within 60 days of their index test. Taken together, our findings suggest that early receipt of nirmatrelvir-ritonavir within 0–5 days after symptom onset likely best reduces risk of hospital admission or death for cases testing positive for SARS-CoV-2 in outpatient settings, underscoring the continued need for prompt testing and treatment among cases at high risk for progression to severe COVID-19.

Postlicensure studies of nirmatrelvir-ritonavir effectiveness in Israel, Hong Kong, and the United States have yielded discordant results, estimating 21–79% effectiveness against severe disease or hospital admission endpoints in outpatient samples.^8–14^ Differences in the populations enrolled, as well as the circumstances of nirmatrelvir-ritonavir access and availability within each setting, merit consideration when reconciling these findings. Studies undertaken during the initial rollout of nirmatrelvir-ritonavir generally enrolled populations at high risk of disease progression; for instance, in two studies in Israel, the mean age of treated cases was 67–69 years, and the prevalence of obesity, diabetes, hypertension, and cardiovascular disease among treated cases each exceeded 30%.^8,10^ One of these studies estimated greater effectiveness of nirmatrelvir-ritonavir among adults aged ≥65 years than adults aged 40–64 years, and among adults who had received ≥2 doses or experienced prior natural infection in comparison to those without previous immunity.^8^ However, such patterns are inconsistent across reports. One US study estimated greater effectiveness among adults aged <65 years than those aged 65–79 or ≥80 years, while another US study^14^ and a separate study in Hong Kong^12^ did not identify differences in effectiveness across subgroups defined by age, immunity, or presence of comorbidities. In further contrast to findings in Israel, studies in Hong Kong^11,12^ which have enrolled the most elderly samples (majority of treated cases aged >70 years) with the lowest vaccine coverage (33–42% considered fully vaccinated against COVID-19) have generated the lowest estimates of effectiveness of nirmatrelvir-ritonavir, spanning 21–33%. Differences across settings or over time in hospital admission criteria for COVID-19 cases may also contribute to variation in effectiveness estimates.

Because the perceived risk for a cases to progress to severe disease likely factors into clinical decision-making around nirmatrelvir-ritonavir prescribing and cases’ adherence to treatment, controlling for potential differences in clinical status among cases who receive or do not receive treatment is important for causal inference in pharmacoepidemiologic studies. Whereas we identify time from symptom onset as a potential modifier of treatment effectiveness, symptoms data were lacking in prior observational studies from all^8–12,14^ or most^13^ cases analyzed. Our estimate of 53.6% effectiveness against the primary endpoint for cases treated at any time in their clinical course aligns with estimates of 45–51% effectiveness against hospitalization in other US observational studies which did not collect symptoms data or restrict according to symptom onset timing.^9,14^ Other unique features of KPSC data may also have helped to control for differences in clinical status and healthcare-seeking behavior in our study. These included the opportunity to match cases according to whether they received clinical care in association with testing (a potential proxy for disease severity); comprehensive information on healthcare-seeking behavior from prior-year interactions across outpatient, ED, and inpatient settings; and documentation of receipt of other vaccines, including 2021-22 seasonal influenza vaccination. Whereas cases with higher rates of prior-year healthcare interactions, higher BMI, and greater burden of comorbid conditions were more likely to receive nirmatrelvir-ritonavir in our sample, the strength of these associations varied considerably across age groups (**Table S3**) and over time (**Figure 2**). Matching cases within regression strata provided an opportunity to account for potential interactions among variables associated with cases’ likelihood of receiving treatment, as observed within our sample, as well as potential interactions among these variables influencing cases’ risk for adverse clinical outcomes.^35,36^ Covariate adjustment^8,10,14^ and inverse probability weighting^9,11^ frameworks applied in other studies may have afforded less flexibility as a strategy to mitigate confounding.

Our study has at least eight limitations. First, capture of several variables is incomplete within KPSC data. While multiple imputation provided a principled approach to address missing data on smoking or BMI, potential misclassification of cases’ immunity from undiagnosed prior infections or those never reported to KPSC remains a concern. Second, as our study is observational in nature, unmeasured confounding may hinder causal inference. Regardless, KPSC records provided more comprehensive measures of clinical status and healthcare-seeking behavior than those available in previous studies,^8–14^ yielding a novel opportunity to address factors potentially influencing receipt of nirmatrelvir-ritonavir and disease severity. Third, we cannot verify whether cases who received nirmatrelvir-ritonavir adhered to treatment as recommended. Our findings should thus be interpreted as measuring intention-to-treat effects under real-world conditions of use. Fourth, our approach to variable selection via a directed acyclic graph, and use of matching to accommodate potential interactions among confounding variables, prioritized validity over precision of estimates. Despite this, our analysis retains sufficient power to reject the null hypothesis when estimating effects of the primary exposures on the primary outcome. Fifth, due to relatively lower risk of severe disease within our study highly vaccinated population, primary and secondary endpoints were rarely observed. Concerns about sparse data bias^39^ are nonetheless mitigated by the fact that point estimates across all endpoints, exposures, and subgroups within our study spanned only 43.8% to 92.2%, with this variability largely explained by time between symptom onset and treatment. Our estimates of treatment effectiveness within three or five days of symptom onset closely matched the findings of the EPIC-HR trial.^6^ Sixth, our analysis did not exclude cases who received other antivirals. Molnupiravir was the only other approved antiviral therapy for treatment of mild-to moderate COVID-19 in the outpatient setting during this study period, but was rarely prescribed as its use was reserved for cases with nirmatrelvir-ritonavir treatment contraindications. Seventh, our endpoint of hospital admission or death due to any cause following a positive outpatient SARS-CoV-2 test may have captured admissions unrelated to COVID-19. Although a substantial proportion of SARS-CoV-2 infections among hospitalized patients may be detected due to entry screening, cases’ absolute risk of COVID-19 related hospital admission during SARS-CoV-2 infection greatly exceeds their risk of hospitalization due to other causes within the same timeframe.^40^ The proportion of hospital admissions unrelated to COVID-19 during follow-up after outpatient testing is thus expected to be markedly lower than the proportion of hospital admissions in which SARS-CoV-2 infection is identified incidentally rather than causally.^41^ If present, misclassification of incidental hospital admissions would be expected to bias our results to the null.^42^ Last, aHR estimates should be interpreted in light of potential underlying depletion-of-susceptibles bias. Rapid progression to hospital admission among cases at the greatest risk may contribute to changes over time in the distribution of covariates among treated and untreated cases, confounding the comparison of instantaneous hazards.^30^

In summary, when compared to matched non-recipients, recipients of nirmatrelvir-ritonavir were substantially less likely to experience hospitalization or death following a positive outpatient SARS-CoV-2 test in southern California during the BA.2 and BA.4/BA.5 waves. Early treatment within 0–5 days from symptom onset was associated with greater effectiveness.

## Data Availability

Anonymized data that support the findings of this study may be made available from the investigative team in the following conditions: (1) agreement to collaborate with the study team on all publications, (2) provision of external funding for administrative and investigator time necessary for this collaboration, (3) demonstration that the external investigative team is qualified and has documented evidence of training for human subjects protections, and (4) agreement to abide by the terms outlined in data use agreements between institutions.

## ACKNOWLEDGMENTS

This study was funded by the US Centers for Disease Control & Prevention (grant 75D3-121C11520 to SYT) and the National Institute for Allergy and Infectious Diseases of the US National Institutes of Health (grant R01-AI4812701A1 to JAL).

## Supporting information

**Table S1:**
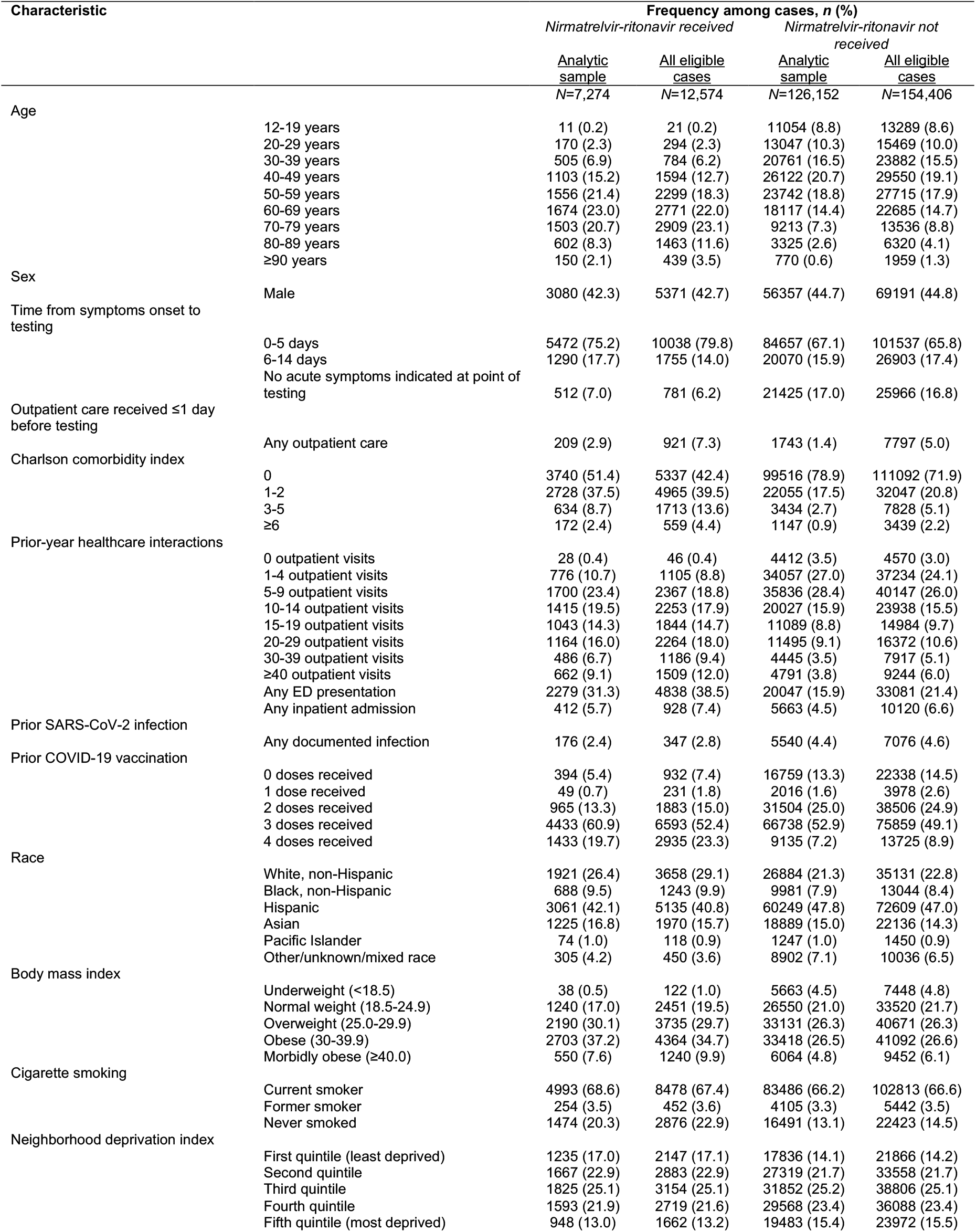
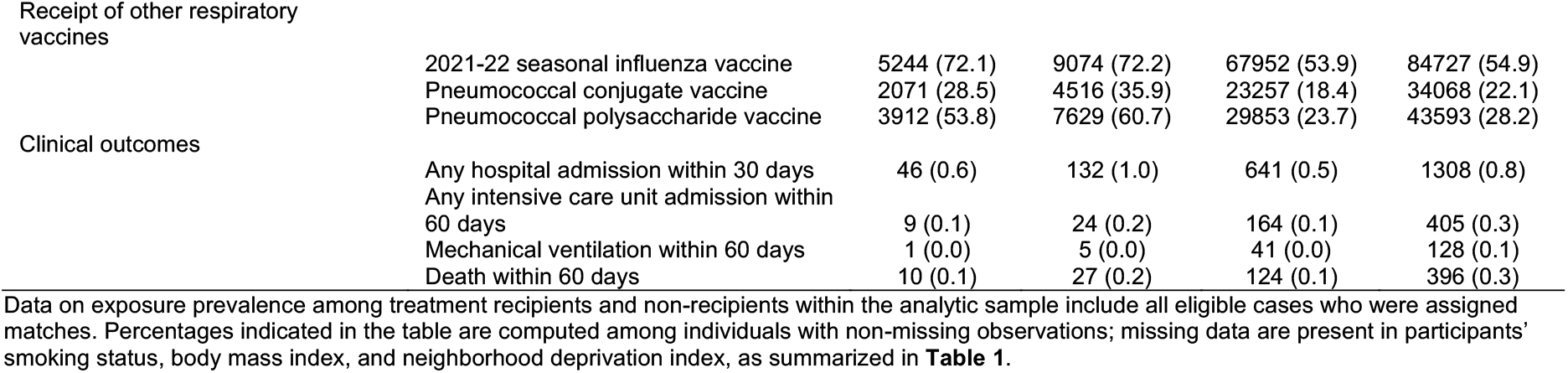
Characteristics of cases receiving and those not receiving nirmatrelvir-ritonavir within the analytic sample and among all eligible cases.

**Table S2:**
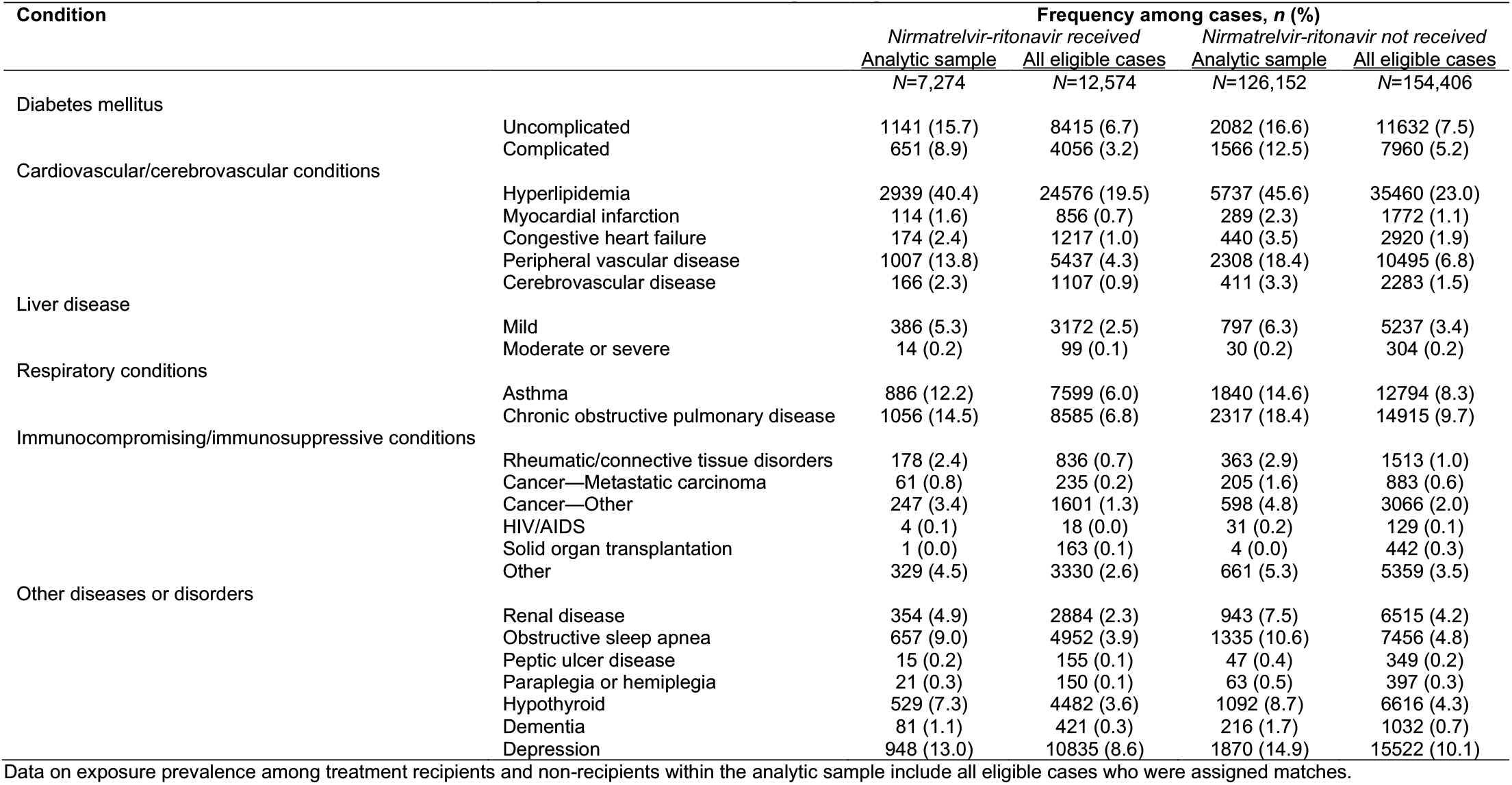
Comorbid conditions within the analytic sample and among all eligible cases.

**Table S3:**
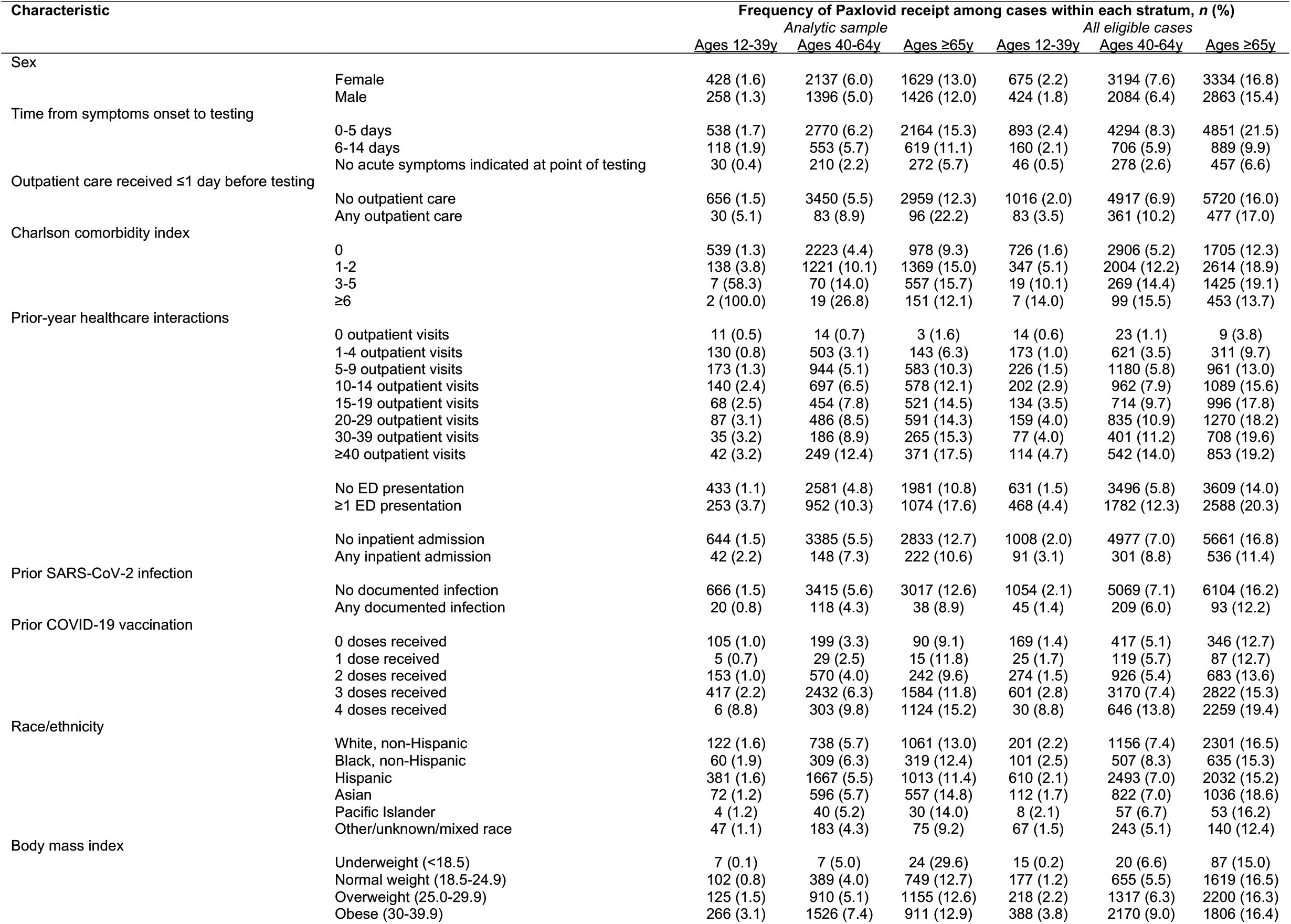
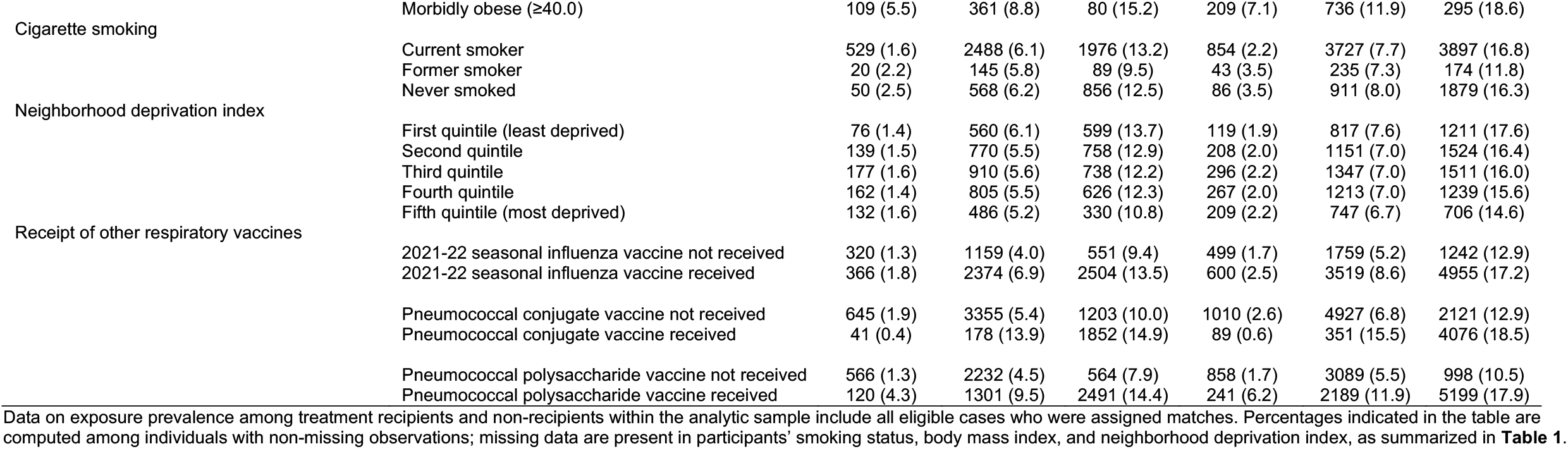
Receipt of nirmatrelvir-ritonavir at ages 12-39 years, 40-64 years, and ≥65 years among cases within the analytic sample and among all eligible cases.

**Table S4:**
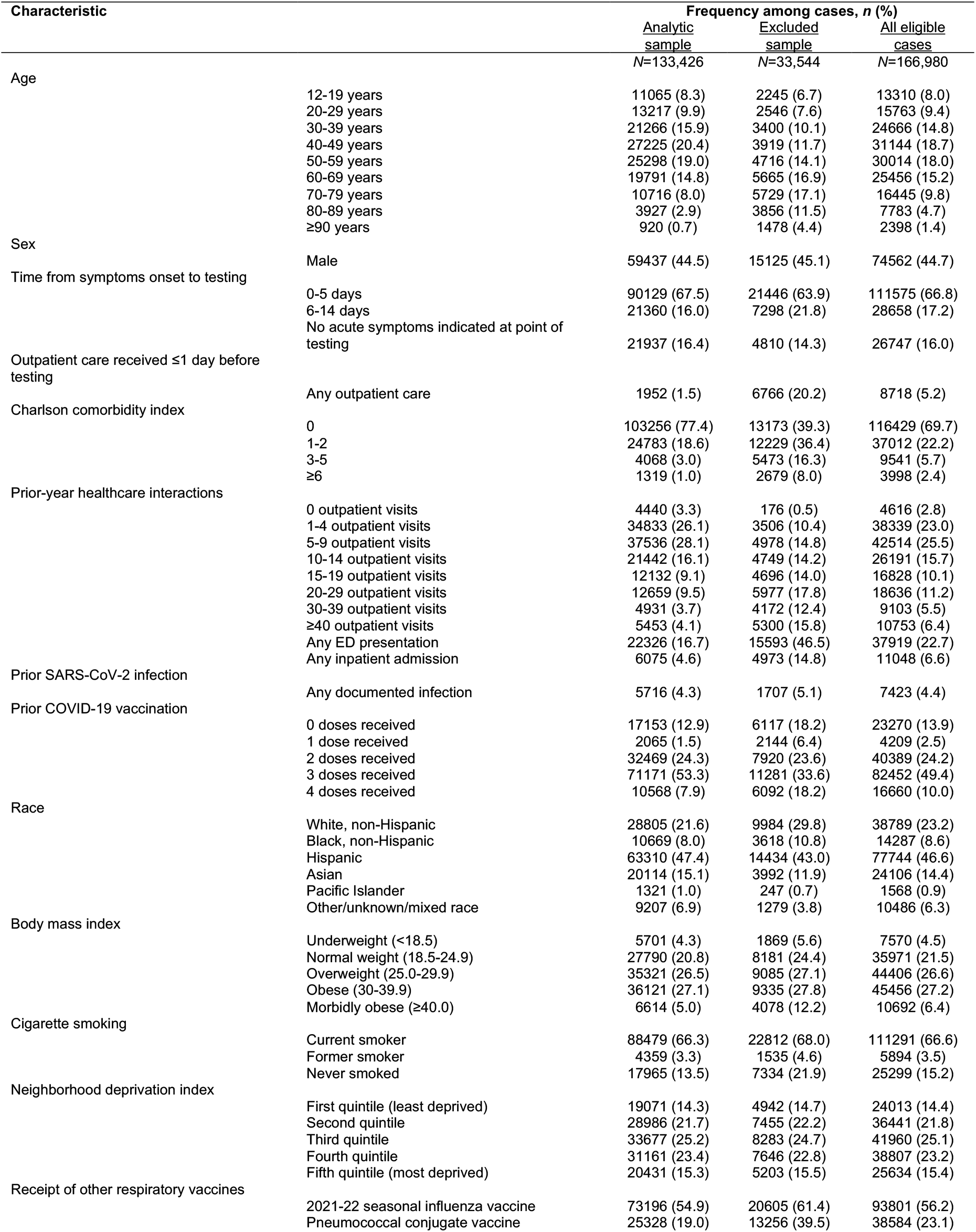
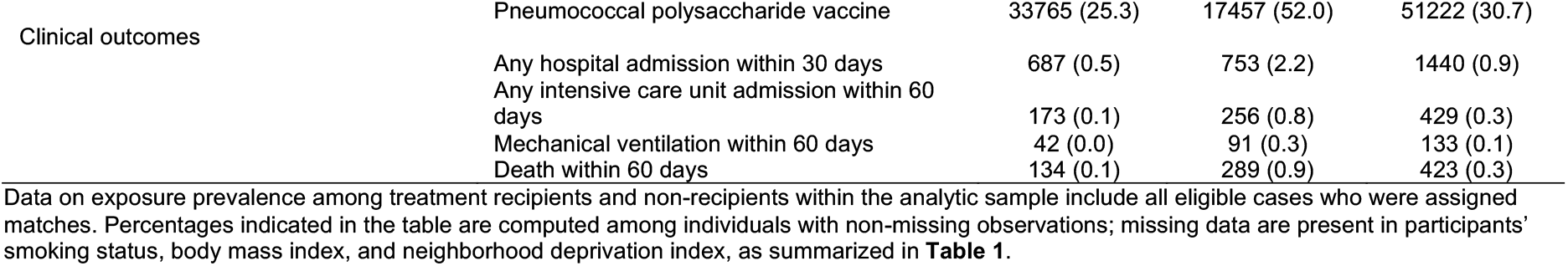
Characteristics of cases included in the analytic sample and those excluded due to a lack of eligible matches.

**Table S5:** Censoring, outcomes, and lengths of follow-up in survival analyses.

| Analytic endpoint | Exposure | Outcomes among treatment recipients |  |  |  | Median follow-up days (IQR) |  |
| --- | --- | --- | --- | --- | --- | --- | --- |
|  |  | Endpoint observed | Change in treatment | Disenrollment or censoring before intended end of follow-up | Censored at intended end of follow-up | Endpoint observed | Endpoint not observed |
| All-cause hospital admission or death within 30 days from positive SARS-CoV-2 test | Nirmatrelvir-ritonavir received | 51 (0.7) | 0 (0.0) | 212 (2.9) | 7,011 (96.4) | 13 (5, 20) | 29 (28, 30) |
|  | Nirmatrelvir-ritonavir not received | 695 (0.5) | 4,725 (3.6) | 2,790 (2.1) | 122,667 (93.7) | 12 (5, 21) | 30 (30, 30) |
| All-cause intensive care unit admission, mechanical ventilation, or death within 60 days from positive SARS-CoV-2 test | Nirmatrelvir-ritonavir received | 17 (0.2) | 0 (0.0) | 1,040 (14.3) | 6,217 (85.5) | 20 (16, 30) | 59 (58, 30) |
|  | Nirmatrelvir-ritonavir not received | 257 (0.2) | 4,725 (3.6) | 14,394 (11.0) | 111,501 (85.2) | 19 (5, 35) | 60 (60, 60) |

Percentages among the arm of cases who did not receive nirmatrelvir-ritonavir are computed from the total number who were not dispensed treatment on their testing date; those who went on to receive treatment at a later point in time are indicated in the "Change in treatment" column, and their outcomes following treatment (censoring or endpoint occurrence) are counted within recipient sample thereafter.

**Table S6:**
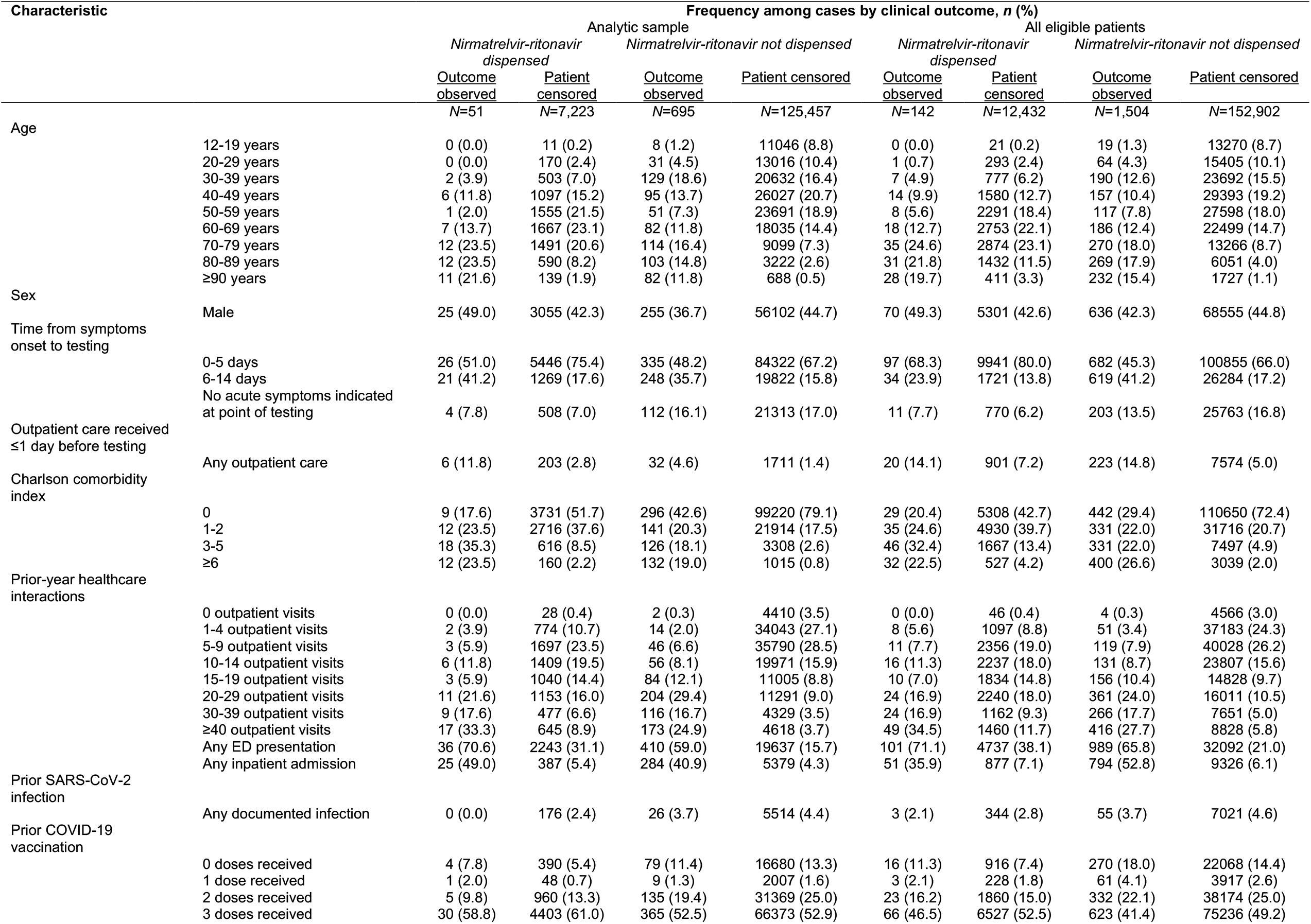
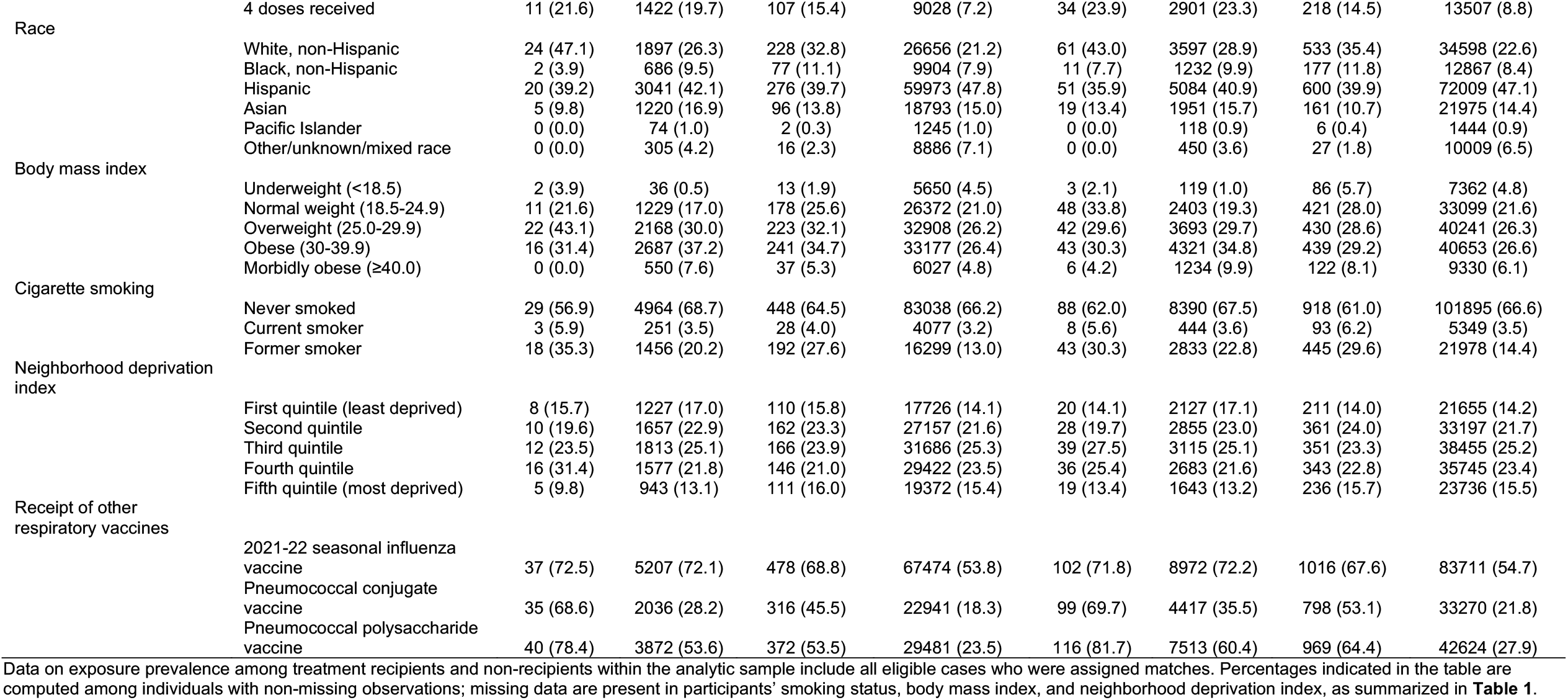
Characteristics of patients experiencing or not experiencing hospital admission or death within 30 days of testing.

**Table S7:**
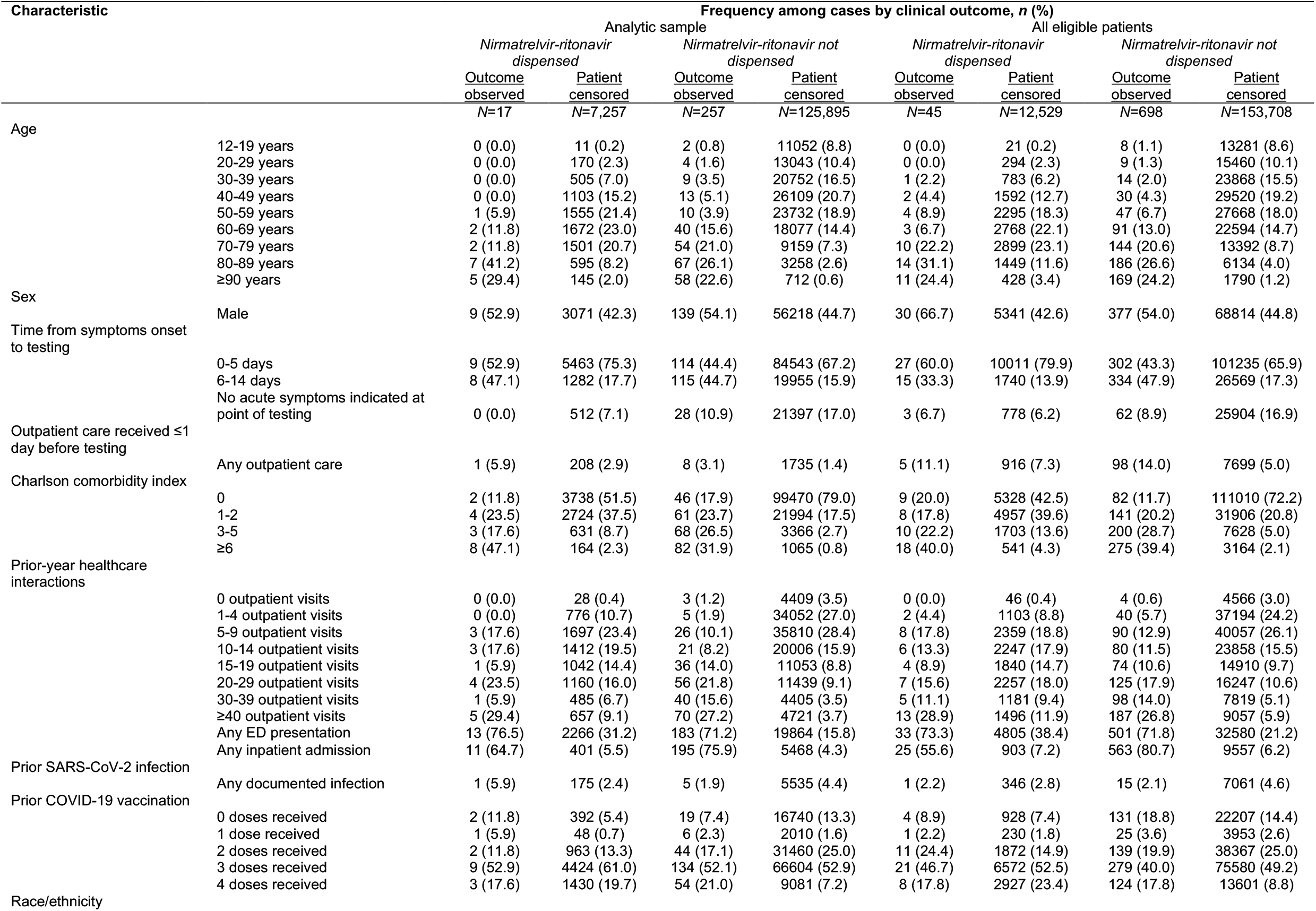
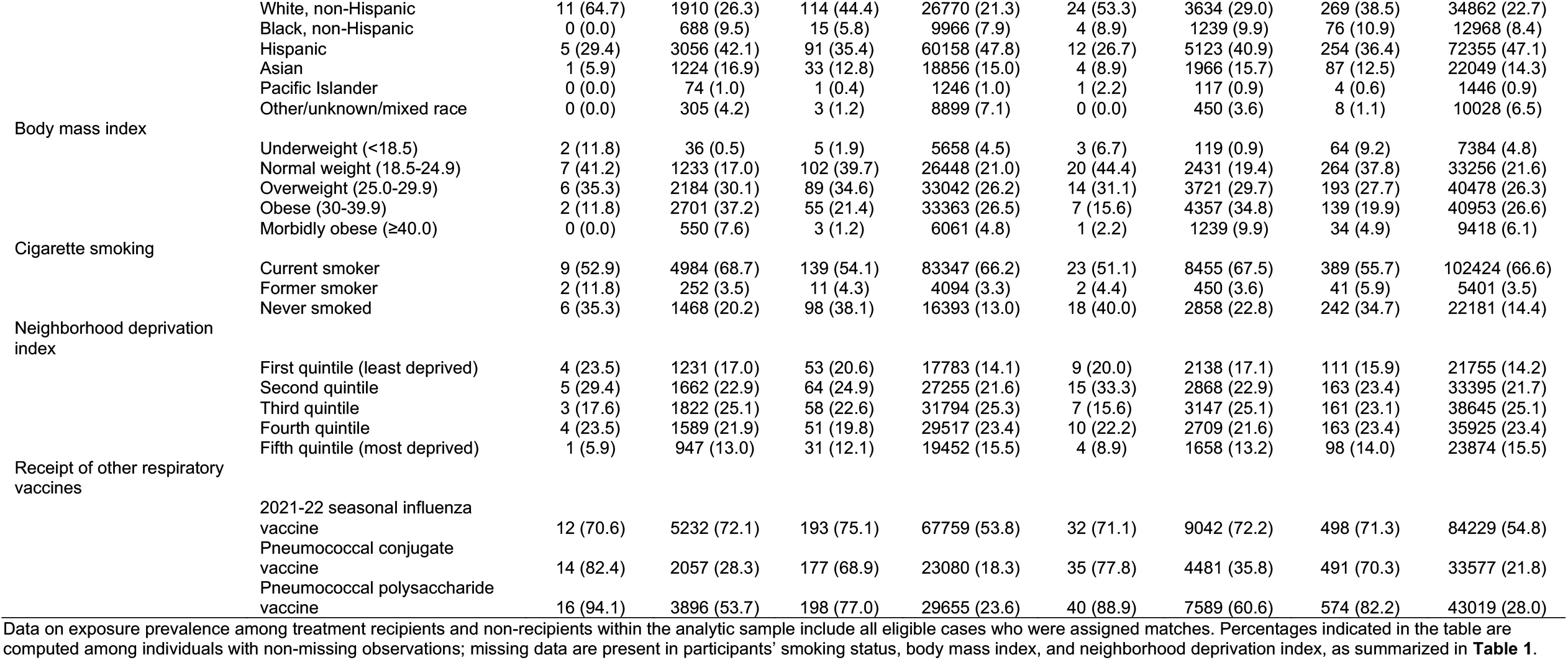
Characteristics of patients experiencing or not experiencing intensive care unit admission, mechanical ventilation, or death within 60 days of testing.

**Table S8:**
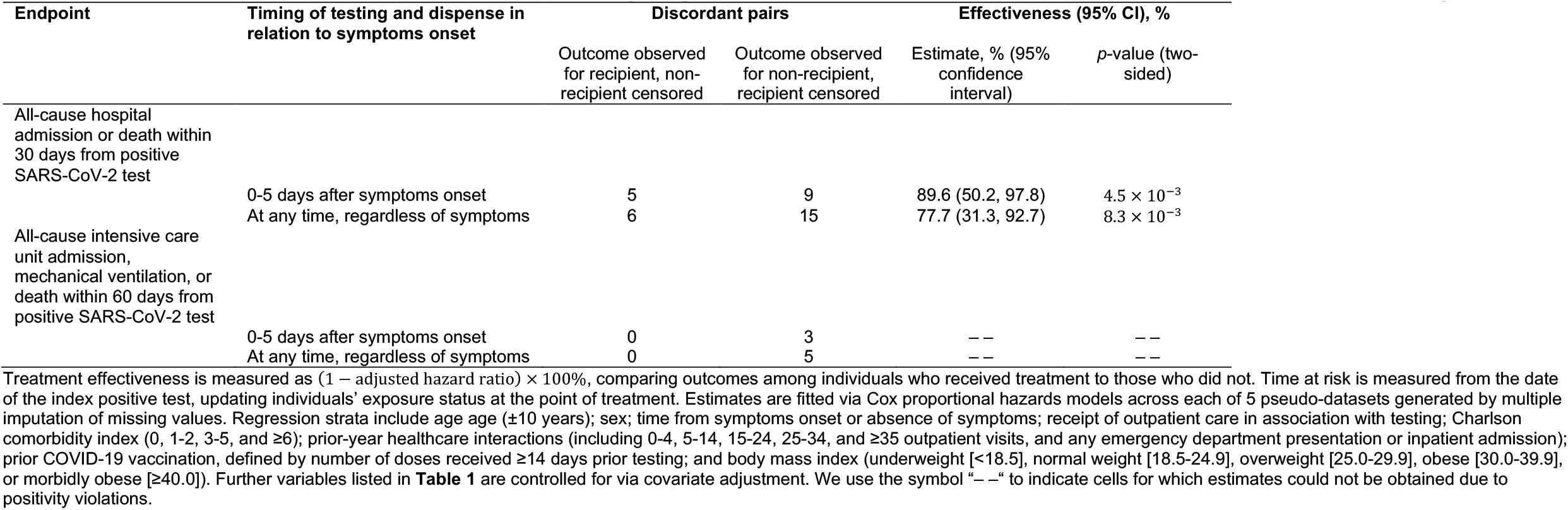
Effectiveness of nirmatrelvir-ritonavir in preventing progression to severe disease endpoints when dispensed on patients’ testing date.

**Table S9:** Effectiveness of nirmatrelvir-ritonavir in preventing progression to severe disease endpoints within subgroups treated at differing stages in the clinical course.

| Endpoint | Timing of dispense in relation to symptoms onset | Discordant pairs |  | Effectiveness |  |
| --- | --- | --- | --- | --- | --- |
|  |  | Outcome observed for recipient, non-recipient censored | Outcome observed for non-recipient, recipient censored | Estimate, % (95% confidence interval) | p-value (two-sided) |
| All-cause hospital admission or death within 30 days from positive SARS-CoV-2 test |  |  |  |  |  |
| | 0-3 days after symptoms onset | 7 | 10 | 81.1 (34.1, 94.6) | $9.0 \times 10^{-3}$ |
|  | ≥6 days after symptoms onset, or while asymptomatic | 18 | 12 | 43.8 (–33.3, 81.7) | 0.16 |
|  | At any time after symptoms onset | 24 | 23 | 59.5 (12.8, 81.2) | 0.021 |
|  | While asymptomatic only | 2 | 0 | – – | – – |
| All-cause intensive care unit admission, mechanical ventilation, or death within 60 days from positive SARS-CoV-2 test |  |  |  |  |  |
|  | 0-3 days after symptoms onset | 1 | 4 | – – | – – |
|  | ≥6 days after symptoms onset, or while asymptomatic | 8 | 4 | 63.6 (–124.6, 94.1) | 0.23 |
|  | At any time after symptoms onset | 10 | 11 | 86.2 (34.6, 97.1) | 0.013 |
|  | While asymptomatic only | 0 | 0 | – – | – – |

**Table S10:**
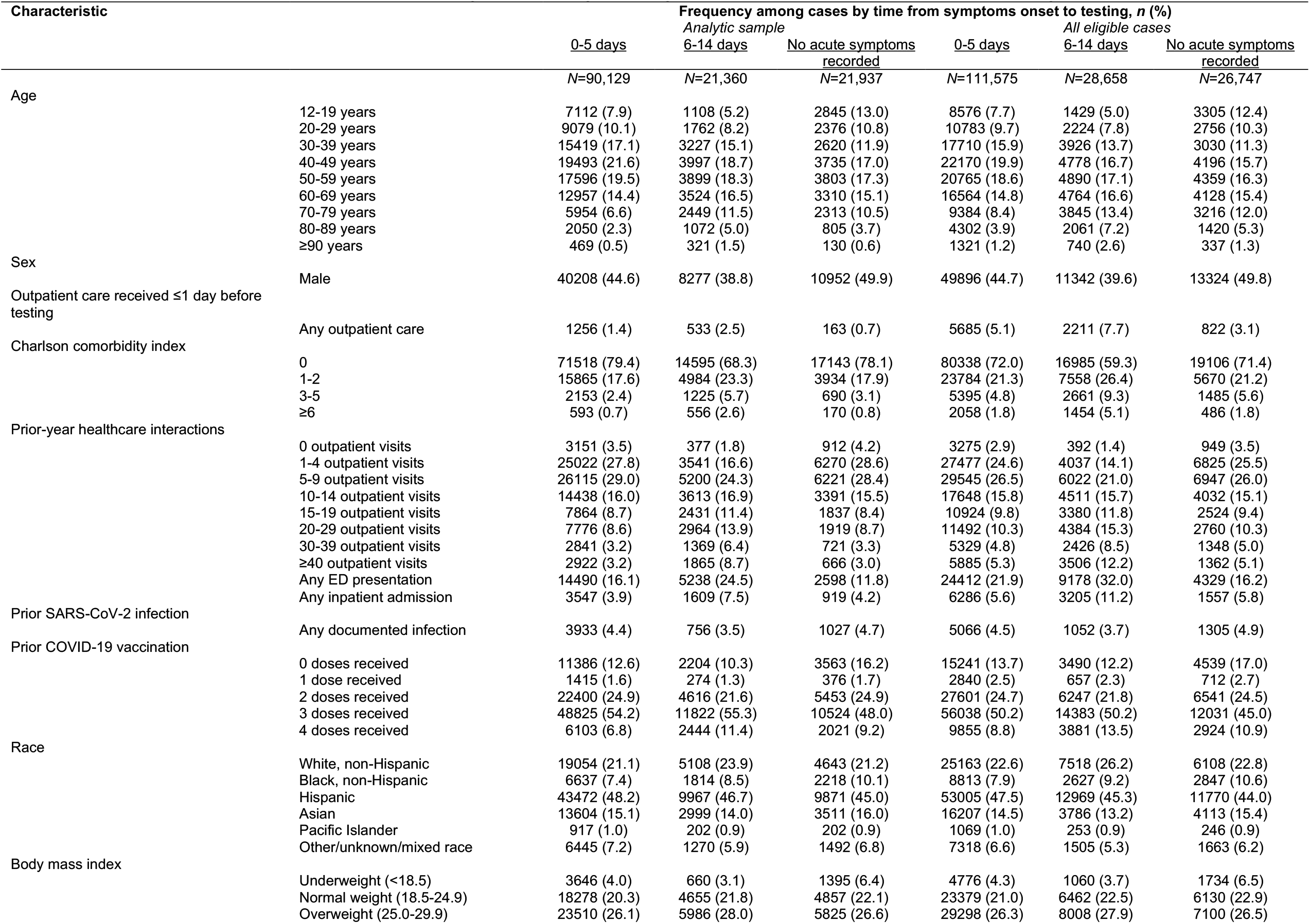
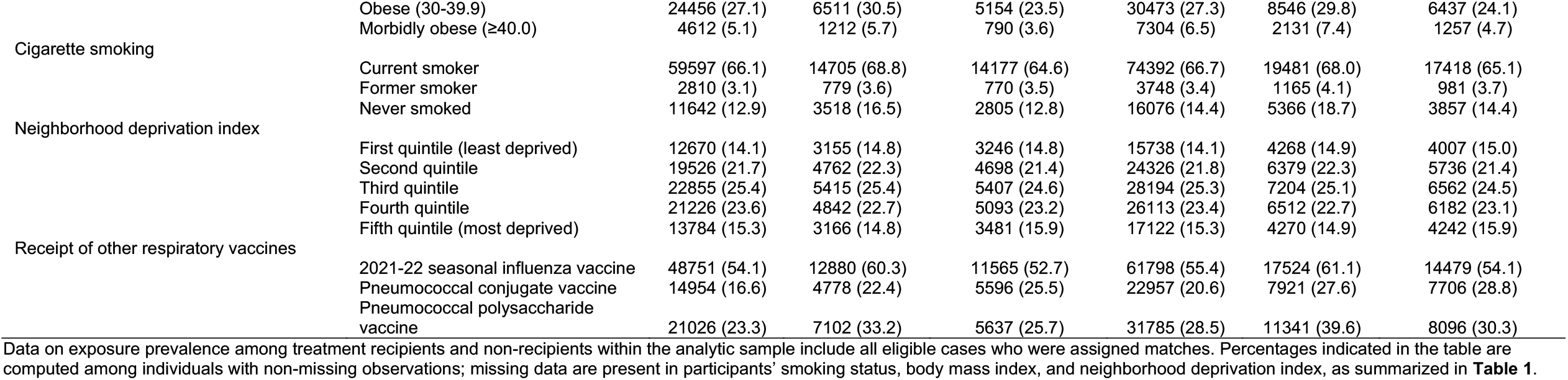
Characteristics of individuals tested 0–5 days or 6–14 days after symptoms onset, or in the absence of acute symptoms.

**Table S11:**
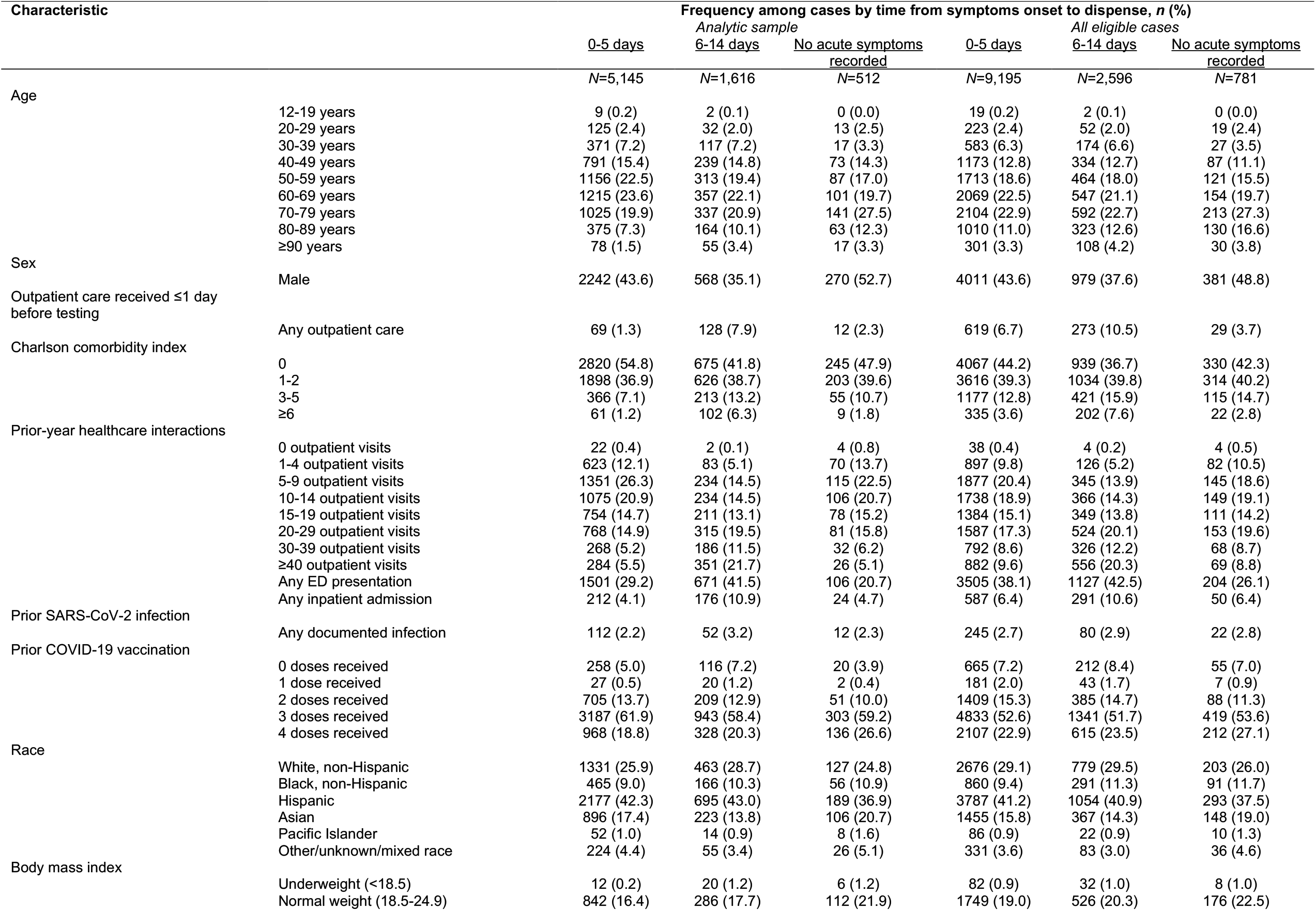
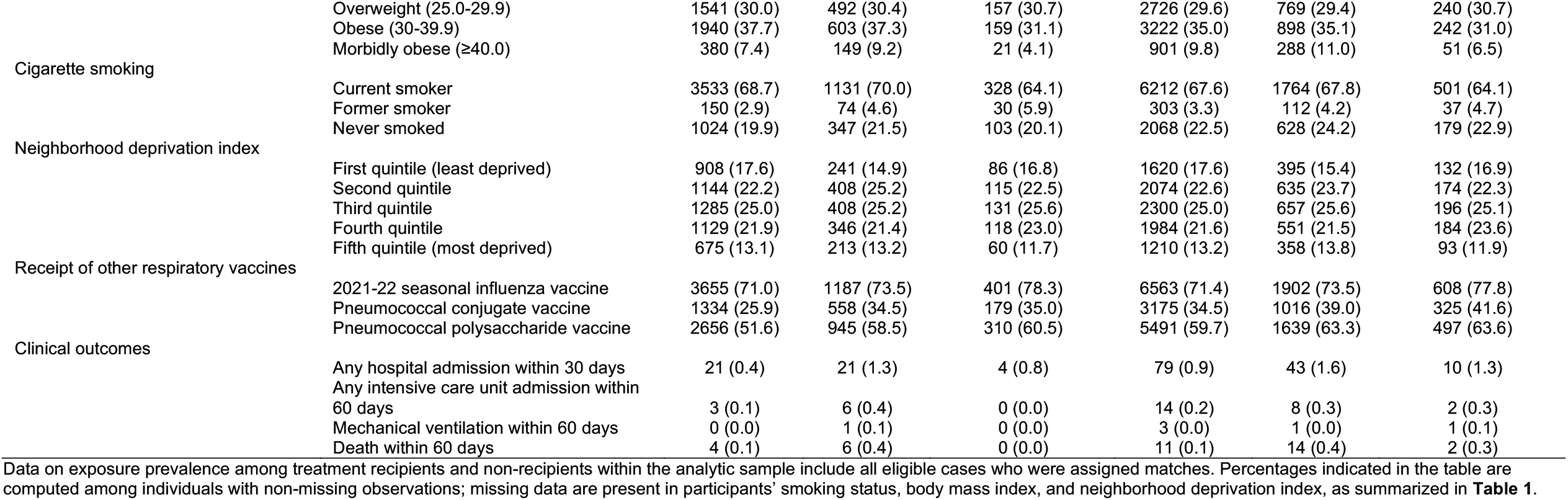
Characteristics of individuals dispensed nirmatrelvir-ritonavir 0–5 days or 6–14 days after symptoms onset, or in the absence of acute symptoms.

**Table S12:**
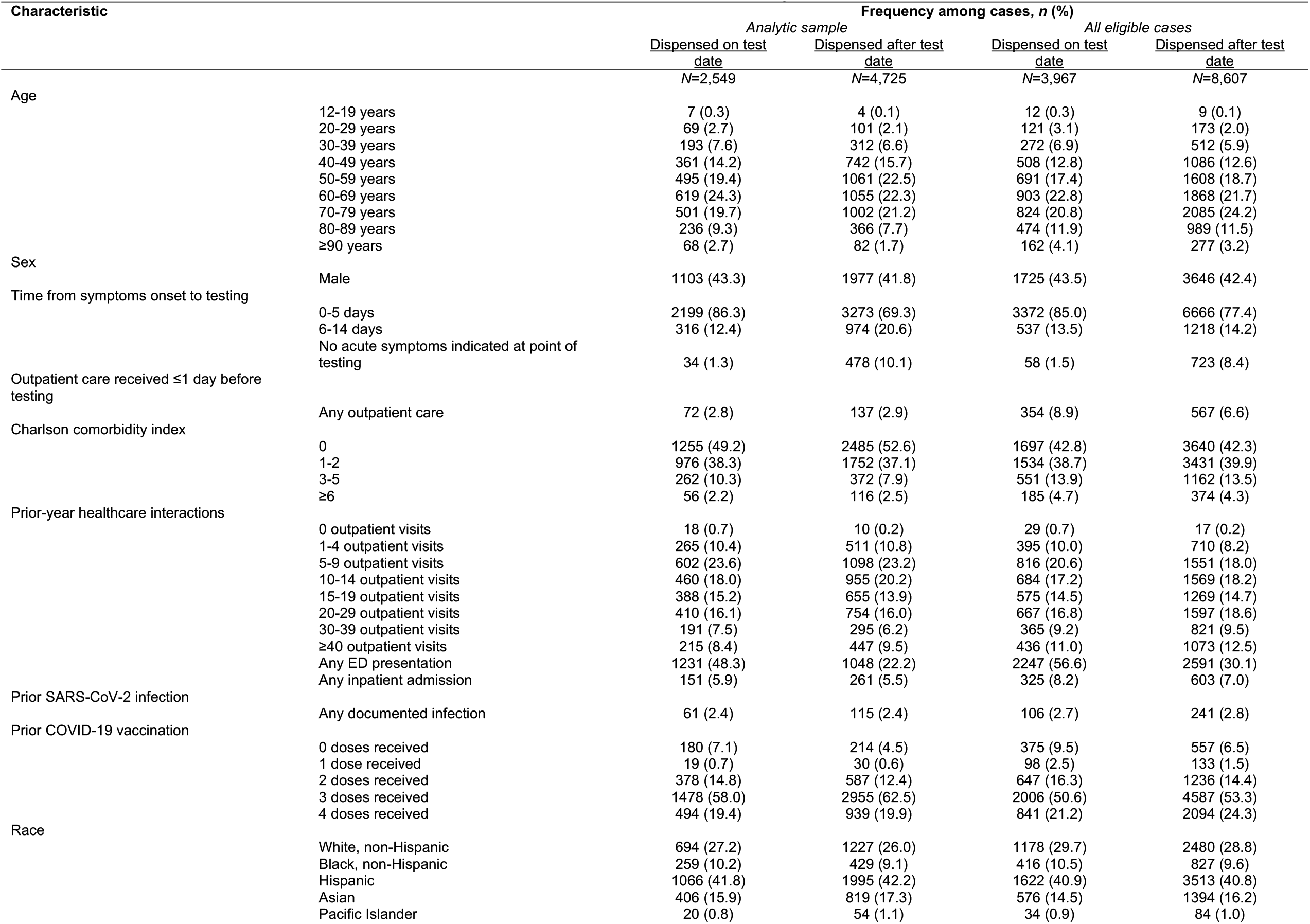
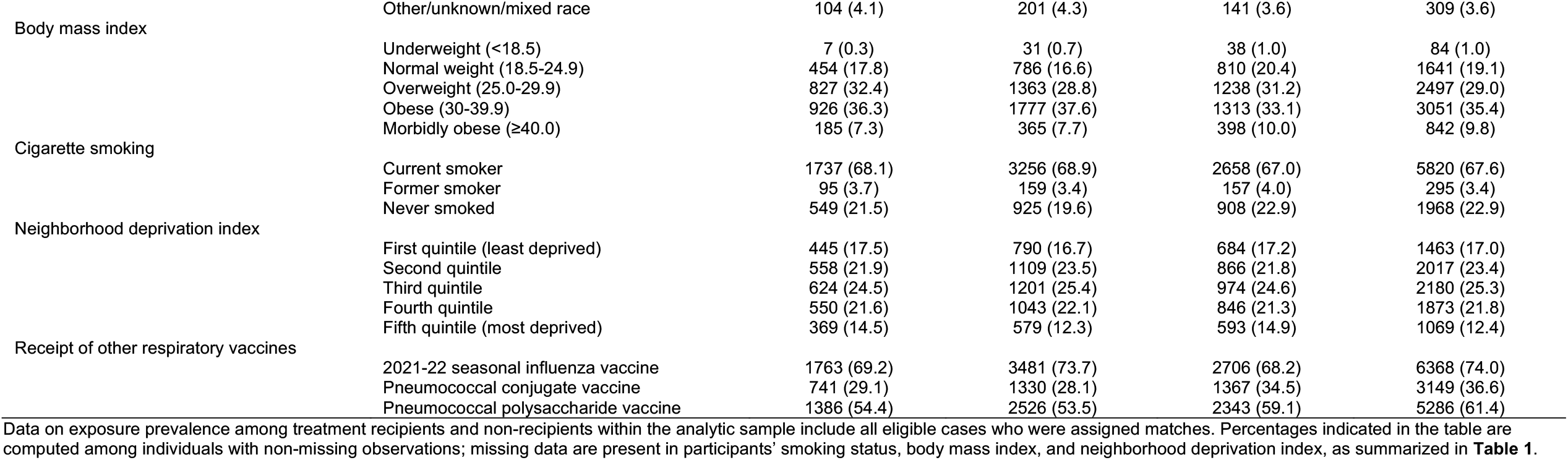
Characteristics of individuals dispensed nirmatrelvir-ritonavir on or after the day of their index test.

**Figure S1:**
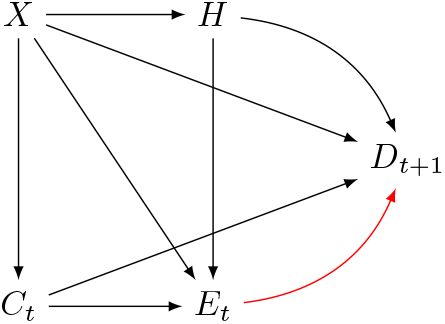
Causal directed acyclic graph. We provide a schematic representation of the causal process used to identify a minimally sufficient set of variables for adjustment. We define the exposure of interest (*E*_*t*_) as receipt of nirmatrelvir-ritonavir at time *t*, and the outcome of interest as severe disease resulting in hospital admission (or other indication of disease progression) at a later point in time, *t*+1, as *D*_*t+*1_. The analytic objective is to measure the effect of *E*_*t*_ on *D*_*t+*1_, plotted as a red arrow. Clinical status at time *t* (*C*_*t*_), risk factors severe illness (*X*), and healthcare-seeking behavior (*H*) jointly influence whether individuals will be exposed to treatment at time *t*. We further hypothesize that the outcome *D*_*t+*1_ is influenced by clinical status at time *t*, patient risk factors, and healthcare-seeking behavior (for instance, if individuals seek pre-hospital care which could prevent or forestall the need for admission at time *t*+1, or if individuals seek and receive hospital care at a given threshold of disease severity). The minimal adjustment set needed to identify the effect of *E*_*t*_ on *D*_*t*+1_ thus includes *X, C*_*t*_, and *H*. We measure *X* via patients’ age, sex, Charlson comorbidity index, history of vaccination and SARS-CoV-2 infection, race, body mass index, cigarette smoking, and neighborhood socioeconomic status. We measure *C*_*t*_ via the presence of symptoms and time from symptoms onset, and by whether individuals received or did not receive clinical care in any outpatient setting in association with testing. We measure *H* via individuals’ prior-year history of healthcare utilization across outpatient, inpatient, and emergency department settings and their receipt of other respiratory vaccines (noting as well that variables such as COVID-19 vaccination, receipt of healthcare in association with testing, and documentation of prior infection may also provide relevant information on *H*, even if their inclusion in the analysis is based on other attributes they are primarily expected to measure).

